# Understanding the impact of high-risk human papillomavirus on oropharyngeal squamous cell carcinomas in Taiwan: A retrospective cohort study

**DOI:** 10.1101/2020.10.26.20213231

**Authors:** Guadalupe Lorenzatti Hiles, Kai-Ping Chang, Emily L. Bellile, Chun-I Wang, Wei-Chen Yen, Christine M. Goudsmit, Hannah L. Briggs, Trey B. Thomas, Lila Peters, Macy A. Afsari, Lisa M. Pinatti, Anna C. Morris, Nadine Jawad, Thomas E. Carey, Heather M. Walline

## Abstract

**Background and Objectives:** Human papillomavirus (HPV)-driven oropharyngeal squamous cell carcinoma (OPSCC) is increasing globally. In Taiwan, HPV-positive OPSCC is obscured by tobacco, alcohol, and betel quid use. We investigated the role of high-risk HPV (hrHPV) in a large retrospective Taiwan OPSCC cohort.

**Methods and Results:** The cohort of 541 OPSCCs treated at Chang Gung Memorial Hospital from 1998-2016 consisted of 507 men (94%) and 34 women (6%). Most used tobacco (81%), alcohol (51%), and betel quid (65%). Formalin-fixed, paraffin-embedded tissue was used for p16 staining (a surrogate marker for HPV) and testing for HPV DNA presence and type by Multiplex HPV PCR-MassArray. HPV DNA and/or p16 staining (HPV-positive) was found in 28.4% (150/528) tumors. p16 and HPV DNA were strongly correlated (*F* < 0.0001). HPV16 was present in 82.8%, and HPV58 in 7.5% of HPV-positive tumors. HPV was associated with higher age (55.5 vs. 52.7 years, *p* = 0.004), lower T-stage (*p* = 0.008) better overall survival (OS) (hazard ratio [HR] 0.58 [95% CI 0.42-0.81], *p* = 0.001), and disease-free survival (DFS) (HR 0.54 [95% CI 0.40-0.73], *p* < 0.0001). Alcohol was strongly associated with recurrence and death (OS: HR 2.06 [95% CI 1.54-2.74], *p* < 0.0001; DFS: HR 1.72 [95% CI 1.33-2.24], *p* < 0.0001). OS and DFS in HPV-positive cases decreased for alcohol users (*p* < 0.0001). Obscured by the strong alcohol effect, predictive associations were not found for tobacco or betel quid.

**Conclusions:** As with HPV-positive OPSCC globally, HPV is an increasingly important etiological factor in Taiwanese OPSCC. HPV-positive OPSCC has considerable survival benefit, but that is reduced by alcohol, tobacco, and betel quid use. hrHPV is a cancer risk factor in males and females. Vaccinating both sexes with a multivalent vaccine including HPV58, combined with alcohol and tobacco cessation policies will be effective cancer-prevention public health strategies in Taiwan.

## Introduction

The occurrence of oropharyngeal squamous cell carcinoma (OPSCC) is rapidly increasing in North America and Western Europe, accounting for approximately 100,000 new cases worldwide each year [1-3]. In particular, the incidence of OPSCC has been dramatically rising since 1973, at the point of surpassing 5% annual increment in the United States in 2000 [2, 4, 5]. OPSCC has been traditionally associated with tobacco use and excessive alcohol consumption as primary risk factors [6-17]. However, recent behavioral changes in Western countries have promoted a marked drop in the prevalence of these major risk factors [7, 13, 18, 19]. In contrast, high-risk human papillomavirus (hrHPV), HPV genotypes 16, 18, 31, 33, 35, 39, 45, 51, 52, 56, 58, 59, 66, 68, and 73, has become the leading etiologic factor of OPSCC [2, 20-28].

Since the World Health Organization recognized the causative link between 15 hrHPV genotypes and the occurrence of OPSCC in 2007 [29], hrHPV has been accepted as a principal etiological cause of this cancer [2, 20, 22, 24, 30-34]. HPV-driven OPSCC is markedly on the rise [2, 4, 10, 22-24, 35-37]. In the United States, the estimated proportion of positive cases has increased from 20% in 1990 to >70%, where hrHPV now represents the most common cause of OPSCC [2, 7, 27, 36, 38]. Countries in Western Europe have observed similar trends [7, 13, 36, 37, 39-41]. Interestingly, these changes have been accompanied by an increment in the survival rates for OPSCC [36, 42, 43]. HPV-positive patients have a significantly better response to treatment (radiation therapy and chemotherapy as well as surgery) and a more favorable prognosis than those diagnosed with HPV-negative OPSCC [23, 25, 27, 28, 35, 42, 44-50].

Despite these observations, recent studies from Taiwan suggest that its high OPSCC rates continue to increase predominantly due to heavy alcohol drinking, cigarette smoking, and betel quid chewing as etiologic factors [9, 51-54]. The strong influence of these risk habits has limited the search for a viral etiology in this population. There have been a few studies indicating that hrHPV is an emerging risk for head and neck cancer in South-East Asia, with a prevalence of HPV-positive OPSCC reported to be absent or present in 12.6% [55-57] to 34% [57-62] of OPSCCs. In this study, we conducted a retrospective cohort analysis to interrogate the prevalence and significance of HPV-driven OPSCC in tissue samples collected from a single major referral site in Taiwan over a period of 18 years. We evaluated the association between clinical characteristics and traditional risk factors (alcohol, smoking, and betel quid) with HPV-associated OPSCC in Taiwan. HPV results were correlated with risk factor exposure for outcomes and survival analysis.

## Methods

### Case identification and study design

This study was performed on a retrospective cohort of OPSCC cases diagnosed from March 1998 to February 2016 at the Chang Gung Memorial Hospital (CGMH)-Linkou in Taoyuan, Taiwan (Taiwan cohort), as described in Figure 1. CGMH is the largest cancer center and a major referral center in Taiwan. Case selection was not a source of bias as we identified OPSCC tumors with a confirmed primary site in the oropharynx using hospital electronic and pathology records from all patients that underwent curative-intent therapy at this tertiary healthcare center. Patients with unknown primary site (T0 or Tx) were excluded. Primary tumors were biopsied only or resected by surgery and collected for histopathological diagnosis. All patients with available pathology-archived tissue and known clinical records were included in this cohort (Fig 1). In total, 541 OPSCC tumors were retrieved, sectioned, anonymized, and shipped to the University of Michigan for HPV and p16 testing. Five non-squamous cell carcinoma cases were excluded from the study based on their pathological classification and histopathological re-assessment of the submitted sections.

**Fig 1.**
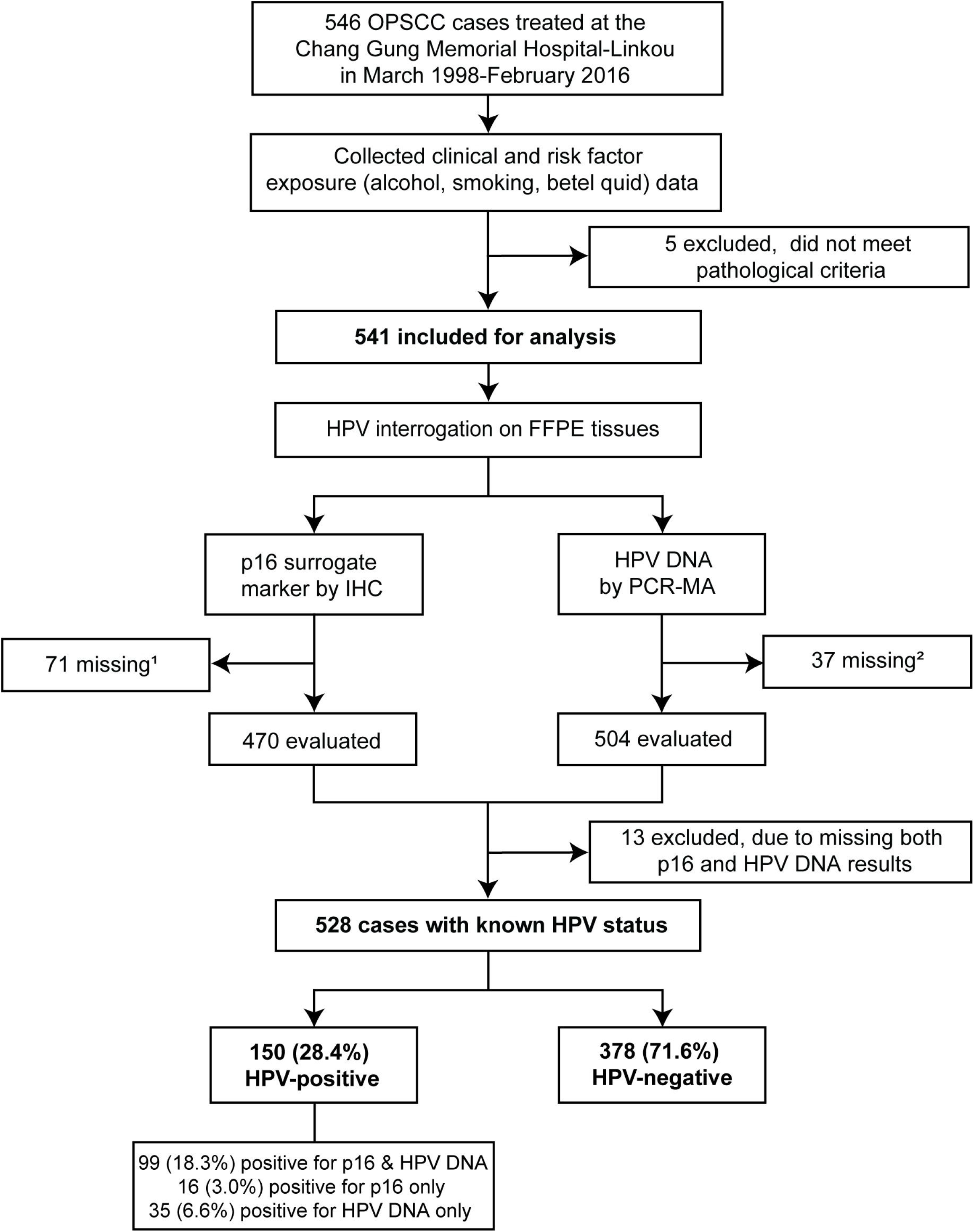
Flow diagram of cases included in the Taiwan retrospective cohort and study design. ^1^Results missing due to absent slide or tissue core, or major artifacts that prevented evaluation. ^2^Results missing due to absent DNA or invalid test. OPSCC, oropharyngeal squamous cell carcinoma; HPV, human papillomavirus; FFPE, formalin-fixed, paraffin-embedded; IHC, immunohistochemical staining; PCR-MA, multiplex PCR-MassArray.

To analyze the contribution of hrHPV on these OPSCC cases, qualitative data on the common risk factors, alcohol consumption, smoking, and betel quid chewing habits were collected. However, because of the retrospective nature of the study, and a change to an electronic record system, data on the quantity of alcohol, smoking, betel quid consumption, or comorbidities could not be retrieved for a large proportion of the patients. Smoking is the only use of tobacco in Taiwan because betel quid preparations do not contain tobacco and tobacco chewing is an extremely uncommon behavior [9, 63]. Demographic information, including patient characteristics (age at diagnosis, and gender), as well as clinical information (stage, tumor site, initial treatment, and outcomes for recurrence, metastasis, and death), were compiled from patient records. Cases were staged at diagnosis according to the seventh edition of the American Joint Cancer Committee (AJCC) [64].

### HPV interrogation

All OPSCC tumors were evaluated for the presence of HPV by two complementary methods: p16 testing and detection of HPV DNA types [65-67] (Fig 1). Results from each determination were blinded to the investigators to avoid bias. Tumors with either p16 and/or HPV DNA positivity were defined as HPV-positive.

#### Detection and genotyping of HPV DNA by Multiplex PCR-MassArray (PCR-MA)

DNA was isolated from tissue curls of formalin-fixed, paraffin-embedded (FFPE) tumor specimens. Two to seven 10-µm FFPE sections were combined in each extraction with AllPrep DNA/RNA FFPE Kit (Catalog No. 80234, QIAGEN, Germantown, MD, US), according to the manufacturer’s recommendations. DNA was eluted in DNase-free water and stored at −20 °C until testing. DNA concentration was measured by a Qubit 2.0 Fluorometer (Catalog No. Q32866, Invitrogen-Thermo Fisher Scientific Inc., Waltham, MA, US) and the Qubit dsDNA HS Assay Kit (Catalog No. Q32851, Invitrogen-Thermo Fisher Scientific Inc., US).

Samples were examined for the presence of HPV DNA and genotyped by PCR-MA analysis, a very sensitive, high-throughput method based on competitive PCR and probe-specific single-base extension coupled with MALDI-TOF mass spectrometry [65-67]. The PCR-MA assay is designed to detect 15 high-risk (HPV 16, 18, 31, 33, 35, 39, 45, 51, 52, 56, 58, 59, 66, 68, and 73), and 2 low-risk (HPV 6 and 11) HPV types, and a possible high-risk subtype (HPV90), as previously described by our laboratory [65-67]. Reactions were prepared with 20 ng of DNA and carried out in quadruplicates in an area physically separated from DNA isolation. Tests were run using a Mass Array 384-format System (Agena Bioscience Inc., San Diego, CA, US). Specimen acceptability was determined using human glyceraldehyde-3-phosphate dehydrogenase (GAPDH) DNA control.

#### p16 testing by immunohistological analysis

FFPE tissue sections (4-µm) were used for p16 immunostaining with a specific antibody against Protein Cyclin-dependent kinase inhibitor 2A (CDKN2A), also known as p16INK4a, as a surrogate for transcriptionally and translationally active HPV [68]. The immunohistochemical (IHC) staining was carried out manually using the clinically validated CINtec-p16 (E6H4) antibody (pre-diluted, Ref. No. 725-4713, Ventana Medical Systems Inc., Tucson, AZ, USA) as stated by the supplier’s protocol. The CINtec-p16 primary antibody was incubated for 1 hour at room temperature followed by washing and appropriate horseradish peroxidase-labeled secondary antibody for 30 minutes at room temperature. All slides were stained with 3,3′-diaminobenzidine for 1-5 minutes, followed by hematoxylin counterstain.

p16 IHC was examined for each slide at 200x and 400x magnification according to the 2018 recommendations of the College of American Pathologists [68]. p16 expression was scored as positive if ≥ 70% of the tumor cells exhibited strong and diffuse nuclear and cytoplasmic p16 immunoreactivity (S1 Fig).

### Statistical analysis

Two-sided Fisher’s exact test (*F*) was used to analyze the relationship between p16 and HPV DNA results. The association between HPV prevalence and study year was evaluated by two-sided simple linear regression and Spearman rank correlation (ρ). Data for HPV status, alcohol consumption, cigarette smoking, betel quid chewing, age, gender, T-stage, N-stage, disease site, initial treatment, and clinical outcomes were collected as covariates. Standard descriptive statistics were performed for each covariate collected. Differences in the distribution of covariates by HPV status were tested by two-sided t-test (continuous measures) or Pearson’s chi-squared test (χ^2^) (categorical/binary). Time-to-event outcomes were defined beginning from date of pathology diagnosis to death from any cause (Overall Survival), or from date of pathology diagnosis to date of first recurrence or death (Disease-Free Survival); subjects alive with no event were censored at date of last follow-up. The Kaplan-Meier method and log-rank tests were used to estimate survival probabilities and plot survival distributions. Cox proportional hazard models and hazard ratio (HR) estimations for time up to 5 years post-diagnosis were performed to test relative hazards between groups in the whole cohort and in subsets stratified by HPV status or other risk factors (alcohol consumption, cigarette smoking, and/or betel quid chewing), adjusting for age, T- and N-stage. Cases with no HPV status data (p16 or HPV DNA), N = 13, were excluded, leaving 528 cases eligible for Chi-squared and survival calculations. Tests were also performed to examine the variates and survival distributions by gender. Statistical analyses were conducted in SAS v9.4 (SAS Institute Inc., Cary, NC, US) using R v3.6.1 (RStudio, Boston, MA, US) for graph generation, or GraphPad Prism v8.3.0 (GraphPad Software, San Diego, CA, US). Statistical tests were performed using 95% confidence intervals and a 5% significance level.

### Ethics statement

This retrospective study was approved by the Institutional Review Boards of the University of Michigan Medical School and the Chang Gung Memorial Hospital and conducted in compliance with the ethical guidelines of the World Medical Association’s Declaration of Helsinki (1964, amended in 2013) and local regulations. Additional patient consent was not required by the institutional review boards as this OPSCC cohort comprised secondary use of tissue specimens with unidentified chart data. All information stripped of personal identifiers to ensure that the data cannot be linked to individual cases in this cohort, are available in the supplementary S1 Table. The procedures described in this manuscript followed the reporting standards for human subject research of the EQUATOR Network, which are detailed in the STROBE report for this study (S1 Checklist).

## Results

### HPV status and clinical characteristics

A total of 546 OPSCC cases were obtained from an unbiased retrospective chart review of individuals treated with standard of care therapy from March 1998 to February 2016 at the CGMH in Taiwan (Taiwan cohort). Among these cases, five were not OPSCC according to the pathology records and slide review; these were excluded as they did not fulfill our inclusion criteria. Therefore, the final study cohort included 541 OPSCC cases (Fig 1, S1 Table).

The presence of HPV in FFPE tumor sections of oropharyngeal cancer was assessed by IHC staining for p16, a surrogate marker for HPV [68] (Fig 1, S1 Fig, S1 Table). HPV genotypes were identified by PCR-MA [65-67] using tumor genomic DNA (Fig 1, S1 Table). Of the 541 OPSCC tumors tested, p16 was positive in 115 (21.3%), negative in 355 (65.6%), and 71 (13.1%) could not be scored. HPV detection and genotyping showed that 134 (24.8%) tumors were HPV-positive, 379 (68.4%) HPV-negative, and 37 (6.8%) had insufficient DNA (failed to amplify the GAPDH control) (Fig 1, S1 and S2 Tables). Of the 134 positives, HPV16 was found alone in 103 (76.9%) and HPV58 was the second most frequently found in 10 (7.5%) tumors. HPV66, HPV59, HPV45, HPV39, HPV34, HPV31, HPV18, and HPV6 were also found in 13 (9.7%) individual tumors. HPV16 was also present together with other HPV genotypes (HPV6, HPV18, HPV35, HPV58, or HPV59) in 8 (6.0%) tumors (Table 1, S1 Table). These results indicate, that aside from low-risk HPV6, there are diverse oncogenic HPV genotypes in Taiwan OPSCCs.

**Table 1.** HPV genotypes frequency.

| **HPV Genotype** | **Frequency** | **Percent** | **Cumulative Frequency** | **Cumulative Percent** |
| --- | --- | --- | --- | --- |
| **HPV16** | 103 | 76.87 | 103 | 76.87 |
| **HPV58** | 10 | 7.46 | 113 | 84.33 |
| **HPV35** | 3 | 2.24 | 116 | 86.57 |
| **HPV18** | 2 | 1.49 | 118 | 88.06 |
| **HPV31** | 2 | 1.49 | 120 | 89.55 |
| **HPV45** | 2 | 1.49 | 122 | 91.04 |
| **HPV59** | 2 | 1.49 | 124 | 92.53 |
| **HPV66** | 1 | 0.75 | 125 | 93.28 |
| **HPV6** | 1 | 0.75 | 126 | 94.03 |
| **HPV16 HPV58** | 2 | 1.49 | 128 | 95.52 |
| **HPV16 HPV18** | 2 | 1.49 | 130 | 97.01 |
| **HPV16 HPV6** | 2 | 1.49 | 132 | 98.50 |
| **HPV16 HPV35** | 1 | 0.75 | 133 | 99.25 |
| **HPV16 HPV59** | 1 | 0.75 | 134 | 100 |

Altogether, p16 overexpression was strongly correlated with HPV status in OPSCC, as the concordance between p16 and HPV DNA testing was 94.9% (423 out of 446, *F* < 0.0001, S2 Table). Therefore, for this study, we defined HPV positivity as either positive by p16 or HPV DNA test. Thus, we had 528 OPSCC tumors with HPV status (positive or negative) called. HPV status (p16 and HPV DNA) data were not obtained for thirteen cases and were not included in the analysis (Fig 1, S1 and S2 Tables). The prevalence of HPV-positive OPSCC in the whole cohort was 28.4% (150 out of 528) (Fig 1). Interestingly, when we examined the yearly occurrence of HPV-positive OPSCC, we found that there was a trend for an increase over the 18 years of study, but it failed to reach statistical significance (Fig 2, S3 Table). However, the same trend is significant when we examined the yearly occurrence of p16 alone (Fig. 2, S3 Table). Nonetheless, this result should be carefully interpreted as 71 cases are missing data for p16 (S1-S3 Tables). We also observed a clear increment in the number of HPV-negative cases that drive the growing incidence of OPSCC rates in Taiwan, thereby obscuring the gradual rise of HPV-positive OPSCCs. Nevertheless, our results demonstrate an increasing role of oncogenic HPV and its causal role as an etiologic factor of OPSCC in this population.

**Fig. 2.**
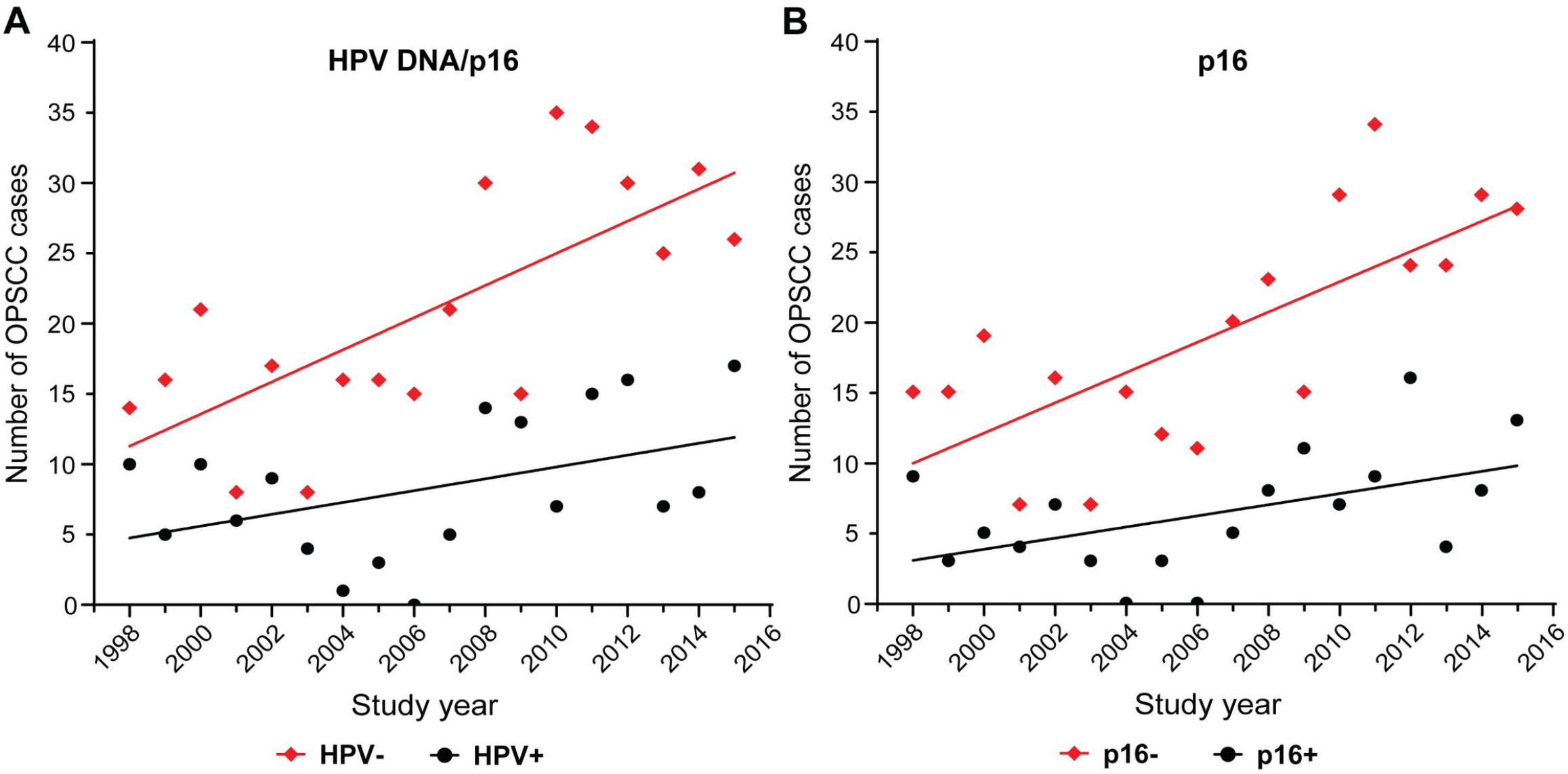
Yearly HPV occurrence among OPSCC cases by HPV DNA and/or p16 (A) or p16 alone (B). The graphs show the correlation between the total frequency of HPV-positive (HPV+) and HPV-negative (HPV-) OPSCC cases, and the study years (see S3 Table). HPV status was assessed by (A) HPV DNA and p16 testing (N = 528) or (B) p16 scoring (N = 458). The association was evaluated in the Taiwan cohort from March 1998 to February 2016 by Spearman’s coefficient (ρ) and linear regression (R^2^). (A) HPV-: ρ = 0.6953, *p* = 0.0014; R^2^ = 0.5201, *p* = 0.0007. HPV+ ρ = 0.4093, *p* = 0.0917; R^2^ = 0.1952, *p* = 0.0664. (B) p16-: ρ = 0.6991, *p* = 0.0012; R^2^ = 0.5555, *p* = 0.0004. p16+ ρ = 0.4741, *p* = 0.0469; R^2^ = 0.2455, *p* = 0.0365.

Next, we assessed the demographic and clinical features of the whole cohort, as listed in Table 2. In our study, males represented 94% (507 out of 541) of all cases, and the average age at tumor diagnosis was 53.5 years old, with 66% of the tumors, diagnosed in individuals of 41-60 years of age (358 out of 541). Over half of the tumors, 58% (315 out of 541), were biopsied or resected from the tonsils, followed by 24% (132 out of 541) from the soft palate, 17% (90 out of 541) from the base of the tongue, and 1% (4 out of 541) from other non-specified locations in the oropharynx. Unknown primary tumors diagnosed by neck node pathology were not included in the tumor retrieval, which may account for the relatively low incidence of base of tongue tumors in this cohort. The large majority of the cohort had a history of previous exposure to known risk factors: 51% (278 out of 541) drank alcohol, 83% (448 out of 541) smoked, and 65% (349 out of 541) chewed betel quid (Table 2, S4 Table). Most prominently, 87% (468 out of 541) of these individuals used more than one of alcohol, and/or tobacco, and/or betel quid concomitantly. Never-smokers, never-drinkers, and never-betel quid chewers accounted for a small 13% (73 out 541) of the cases (S4 Table). Because of the high density of risk factors, and their combined exposure, we were unable to separate their individual effects. Therefore, only 1% (7 out of 541) of the Taiwan cohort was exposed to alcohol without smoking or betel quid chewing, 11% (60 out of 541) solely smoked, and 1% (5 out of 541) only consumed betel quid (S4 Table).

**Table 2.**
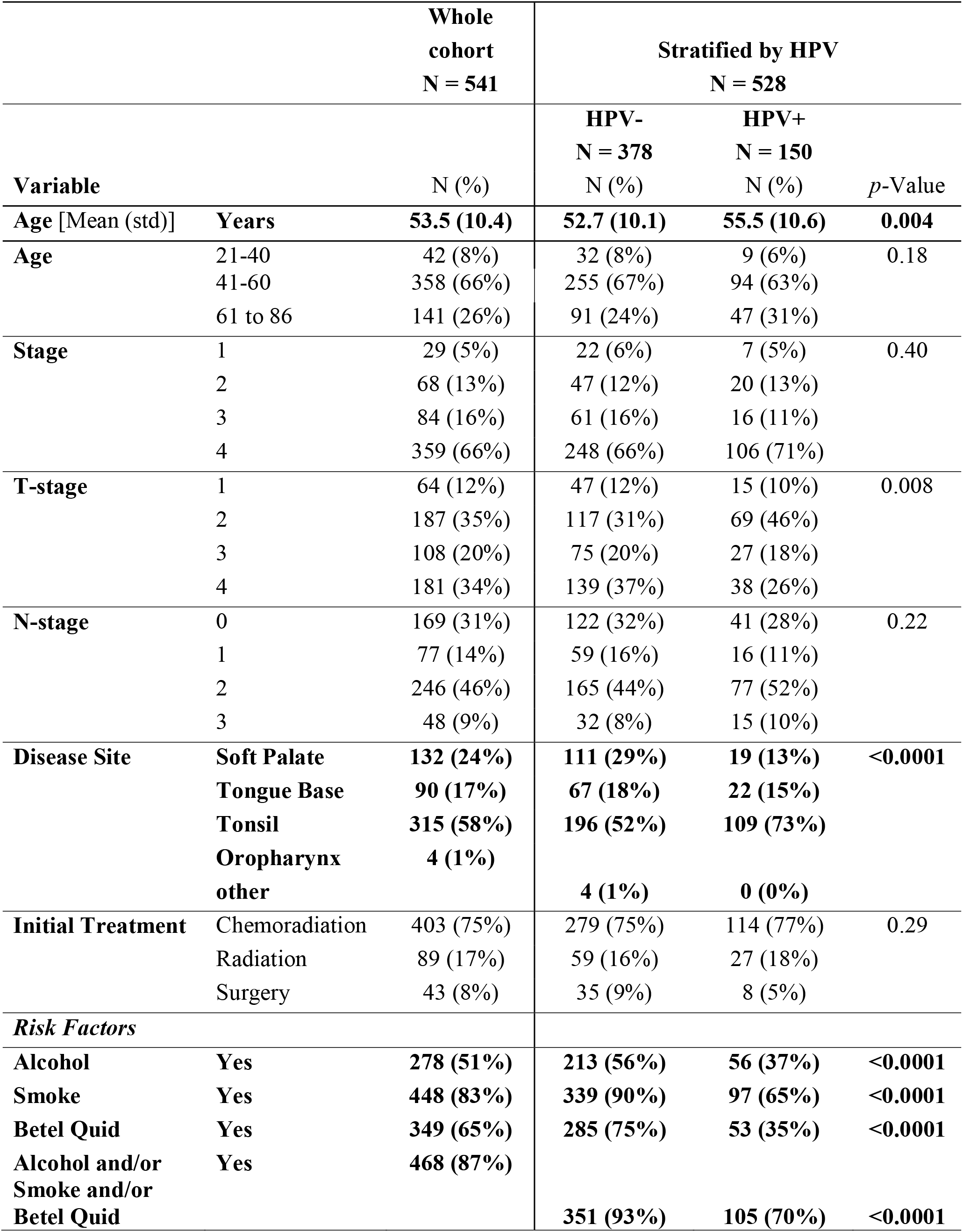
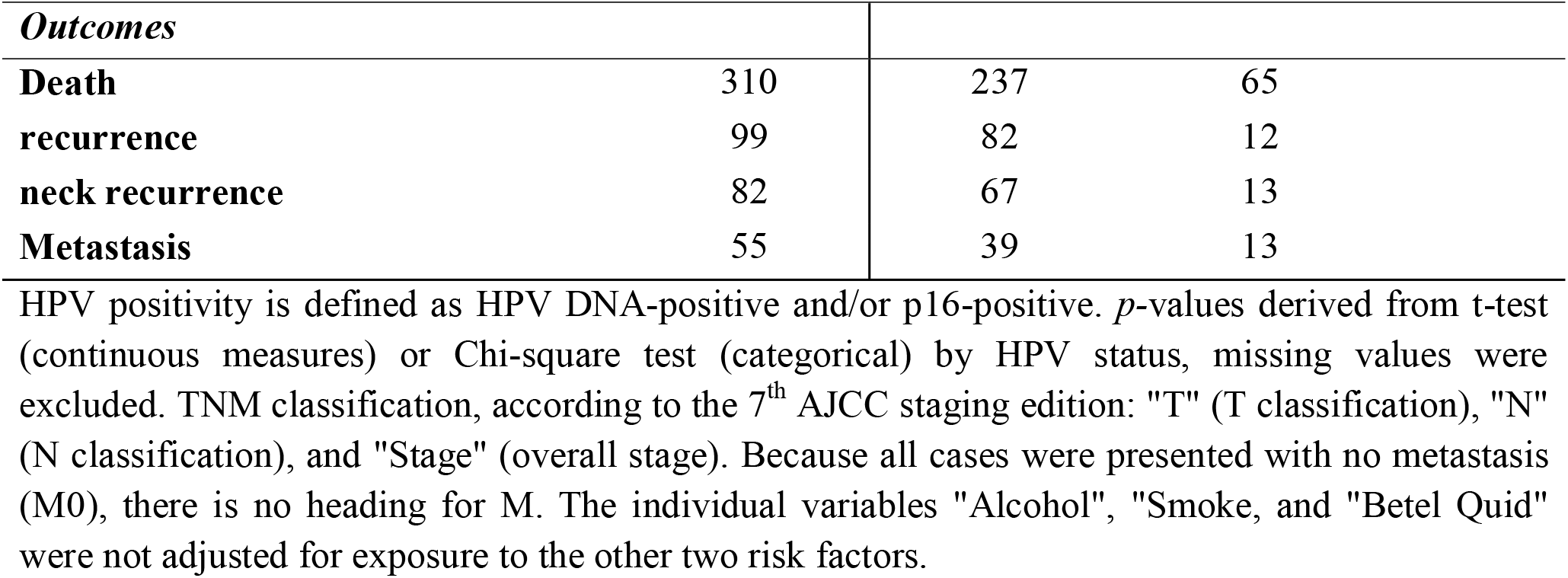
Demographic and clinical characteristics.

We then examined the clinical determinants of HPV status (Table 2). When compared to HPV-negative cases, HPV-positive tumors (HPV DNA-positive and/or p16-positive) had a slightly higher average age at diagnosis (55.5 vs. 52.7 years, *p* = 0.004), with most individuals presenting between 41-60 years old in both groups (67% vs. 63%, *p* = 0.18), and slightly lower T-stage (majority of T2 cases vs. T4, *p* = 0.008). The HPV-positive tumors were also most frequently located in the tonsils (73% vs. 52%, *p* < 0.0001); and presented with lower exposure to all the risk factors including alcohol (37% vs. 56%, *p* < 0.0001), smoking (65% vs. 90%, *p* < 0.0001), and betel quid (35% vs. 75%, *p* < 0.0001) (Table 2, S4 Table). Although the number of females was far lower, representing only 6% (34 out of 541) of the cases (Table 2), they showed pronounced differences with males (S5 Table). The majority, 62% (21 out of 34) of tumors from females, but a minority, 25% (129 out of 507) of tumors from males, were HPV-positive (*p* < 0.0001). The proportion of tonsil tumors was also higher in females than in males (85% vs. 56%, *p* = 0.01), and were more likely to be N3 (*p* = 0.03) (AJCC 7^th^ edition). The females had lower exposure to alcohol (18% vs. 54%, *p* < 0.0001), smoking (21% vs. 87%, *p* < 0.0001), and betel quid (12% vs. 68%, *p* < 0.0001); and were more likely to be never users of these high risk carcinogens. Females also tended to be older at diagnosis (mean: 58.7 years vs. 53.1 years, *p* = 0.002). Among both males and females the majority of cases were of 41-60 years of age (64% v. 66%, *p* = 0.13) (S5 Table). There was also a slightly better prognosis for the female group (OS: log-rank *p* = 0.05; DFS: log-rank *p* = 0.07) (S2 Fig), but given the small group size (N = 34), this observation should be carefully interpreted.

### The role of HPV status and associated risk factors on OPSCC outcome

First, we determined if HPV status had survival benefits on OPSCC by multivariable and Kaplan-Meier analysis (Fig 3, S6 Table). Compared to HPV-negative cases, patients with HPV-positive OPSCC had significantly higher overall survival (HR 0.58, 95% CI 0.42 to 0.81, *p* = 0.001; log-rank *p* < 0.0001) and disease-free survival (HR 0.54, 95% CI 0.40 to 0.73, *p* < 0.0001; log-rank *p* < 0.0001) for up to 5-year post-diagnosis, suggesting that HPV-positivity was an independent predictor for better prognosis.

**Fig 3.**
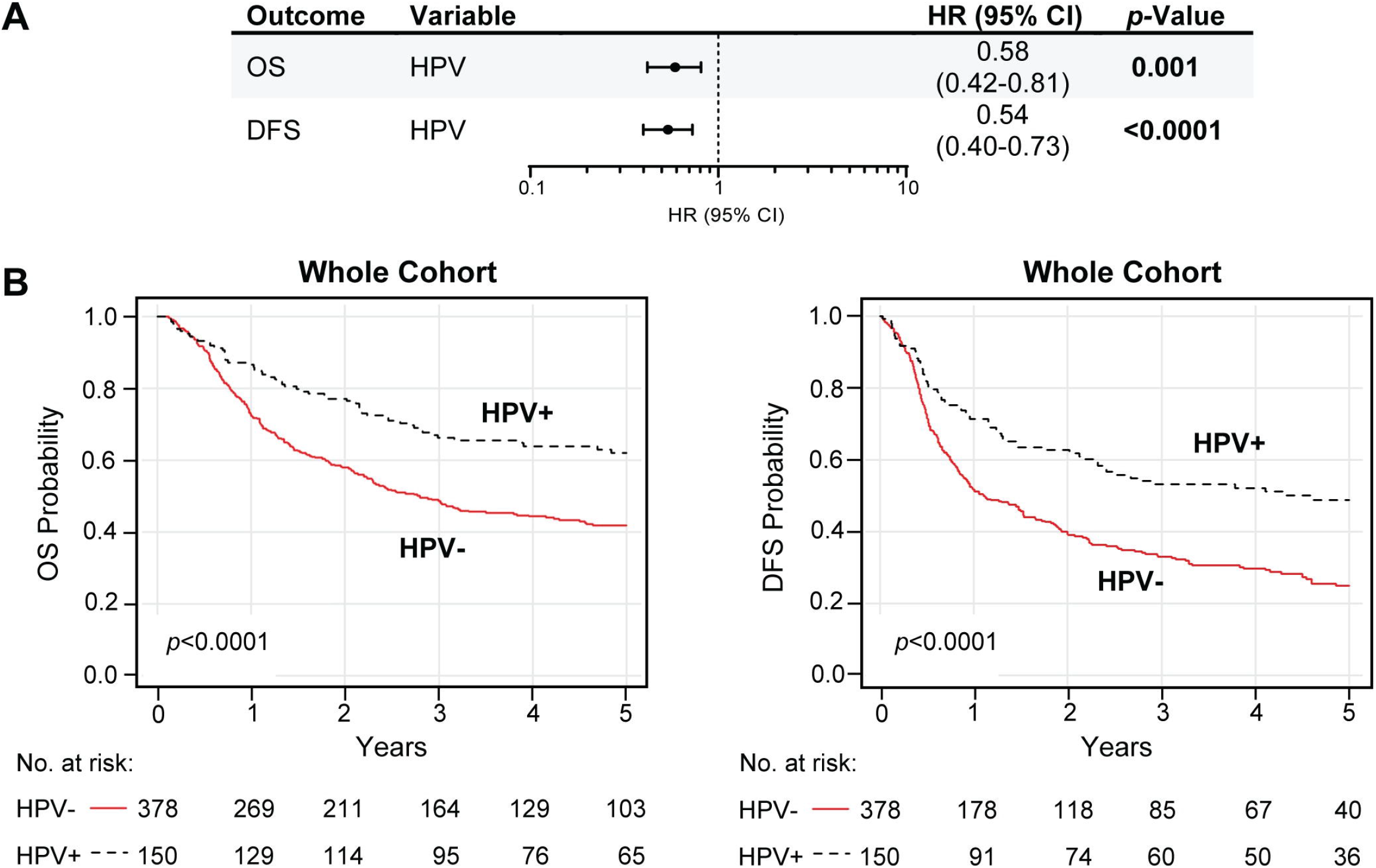
HPV-positive OPSCC is associated with increased survival time. (A-B) Up to 5-year overall survival (OS) and disease-free survival (DFS) prognostic outcomes of the HPV variable in the whole OPSCC Taiwan cohort. HPV positivity is defined as HPV DNA-positive and/or p16-positive. (A) Table includes the multivariable hazard probabilities analyzed using Cox survival models and hazard ratio (HR) estimations, visualized by forest plots. The complete analysis is found in S6 Table, where estimates were reported for full model with all covariates (HPV status, alcohol, smoking, betel quid, age, N- and T-stage) included as fixed effects. (B) Kaplan-Meier survival analysis. Plots represent the results for up to 5-year OS (left) and DFS (right) comparison between HPV-negative (HPV-) and HPV-positive (HPV+) groups. Log-rank analysis was used to compare the survival distributions (log-rank *p*-values are in the plots). HPV-, HPV-negative; HPV+, HPV-positive.

Similar analyses also revealed that consumption of alcohol was a strong negative prognostic factor for both up to 5-year overall survival (OS) (HR 2.06, 95% CI 1.54 to 2.74, *p* < 0.0001; log-rank *p* < 0.0001) and disease-free survival (DFS) (HR 1.72, 95% CI 1.33 to 2.24, *p* < 0.0001; log-rank *p* < 0.0001) (Fig 4, S6 Table). Surprisingly, smoking, and betel quid chewing had no predictive effects (Fig 4, S6 Table). Smoking did not have statistical significance for worse overall survival (HR 0.76, 95% CI 0.50 to 1.14, *p* = 0.18; log-rank *p* = 0.24) and disease-free survival (HR 0.81, 95% CI 0.56 to 1.17, *p* = 0.26; log-rank *p* = 0.40). Betel quid exposure showed no effect on survival by multivariable analysis (OS: HR 0.92, 95% CI 0.67 to 1.27, *p* = 0.60; log-rank *p* < 0.05 – DFS: HR 0.90, 95% CI 0.66 to 1.21, *p* = 0.46; but by Kaplan-Meier analysis betel quid use, just reached significance log-rank *p* < 0.05).

**Fig 4.**
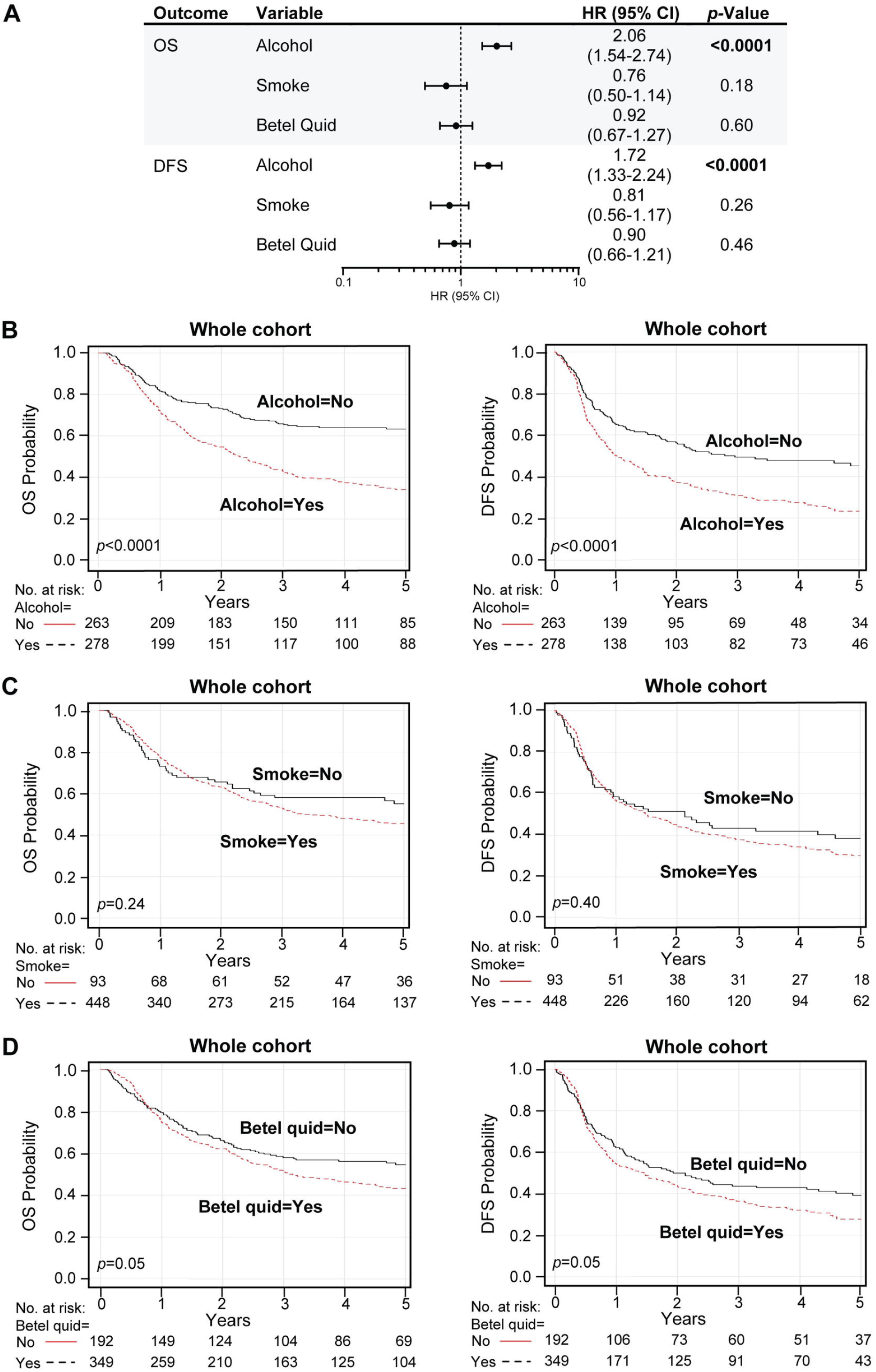
Alcohol is associated with reduced OPSCC survival time. (A-D) Prognostic outcomes of the alcohol, smoking, and betel quid variables within the whole cohort. We analyzed up to 5-year overall survival (OS) and disease-free survival (DFS) outcomes for high-risk habits. (A) Table includes the multivariable hazard probabilities analyzed using Cox survival models and hazard ratio (HR) estimations, visualized by forest plots, where estimates were reported for full model with all covariates (HPV status, alcohol, smoking, betel quid, age, N- and T-stage) included as fixed effects. The complete analysis is found in S6 Table. (B-D) Kaplan-Meier survival analysis. Plots represent the results for up to 5-year OS (left) and DFS (right) comparison between alcohol (B), smoke (C), and betel quid (D) groups. Log-rank analysis was used to compare the survival distributions (log-rank *p*-values are in the plots).

Additional multivariable survival analysis and Kaplan-Meier estimations for alcohol, smoking, and betel quid groups, controlled for HPV status, were also performed (Fig 5, S3 Fig, S7 Table). Alcohol use had an adverse effect on outcome in both HPV-positive and HPV-negative groups, but HPV was associated with longer OPSCC survival time (survival by alcohol within HPV groups, OS: HPV-positive log-rank *p* = 0.0007, HPV-negative long rank *p* < 0.0001 – DFS: HPV-positive log-rank *p* = 0.02, HPV-negative long-rank *p* = 0.0005). No predictive associations were found for tobacco (survival by smoking within HPV groups, OS: HPV-positive log-rank *p* = 0.44, HPV-negative long rank *p* < 0.28 – DFS: HPV-positive log-rank *p* = 0.49, HPV-negative long-rank *p* = 0.18) or betel quid chewing (survival by betel quid within HPV groups, OS: HPV-positive log-rank *p* = 0.41, HPV-negative log rank *p* < 0.68 – DFS: HPV-positive log-rank *p* = 0.21, HPV-negative long-rank *p* = 0.46) based on HPV status. Most importantly, our data demonstrate that the prognostic benefit of HPV positivity persists in the presence of risk factors, alcohol, and tobacco, and betel quid (DFS by HPV within risk groups: alcohol yes log-rank *p* = 0.008, alcohol no log-rank *p* = 0.007; smoking and/or betel quid yes log-rank *p* = 0.0006, smoking and/or betel quid no log rank *p* = 0.01) (S4 Fig, S8 Table). Furthermore, among non-drinkers, non-smokers, and non-betel quid chewers, HPV-positive OPSCC had the best outcomes. In sum, our study shows that hrHPV has a causal role but also carries a significant prognostic benefit for OPSCC in this Taiwan cohort.

**Fig 5.**
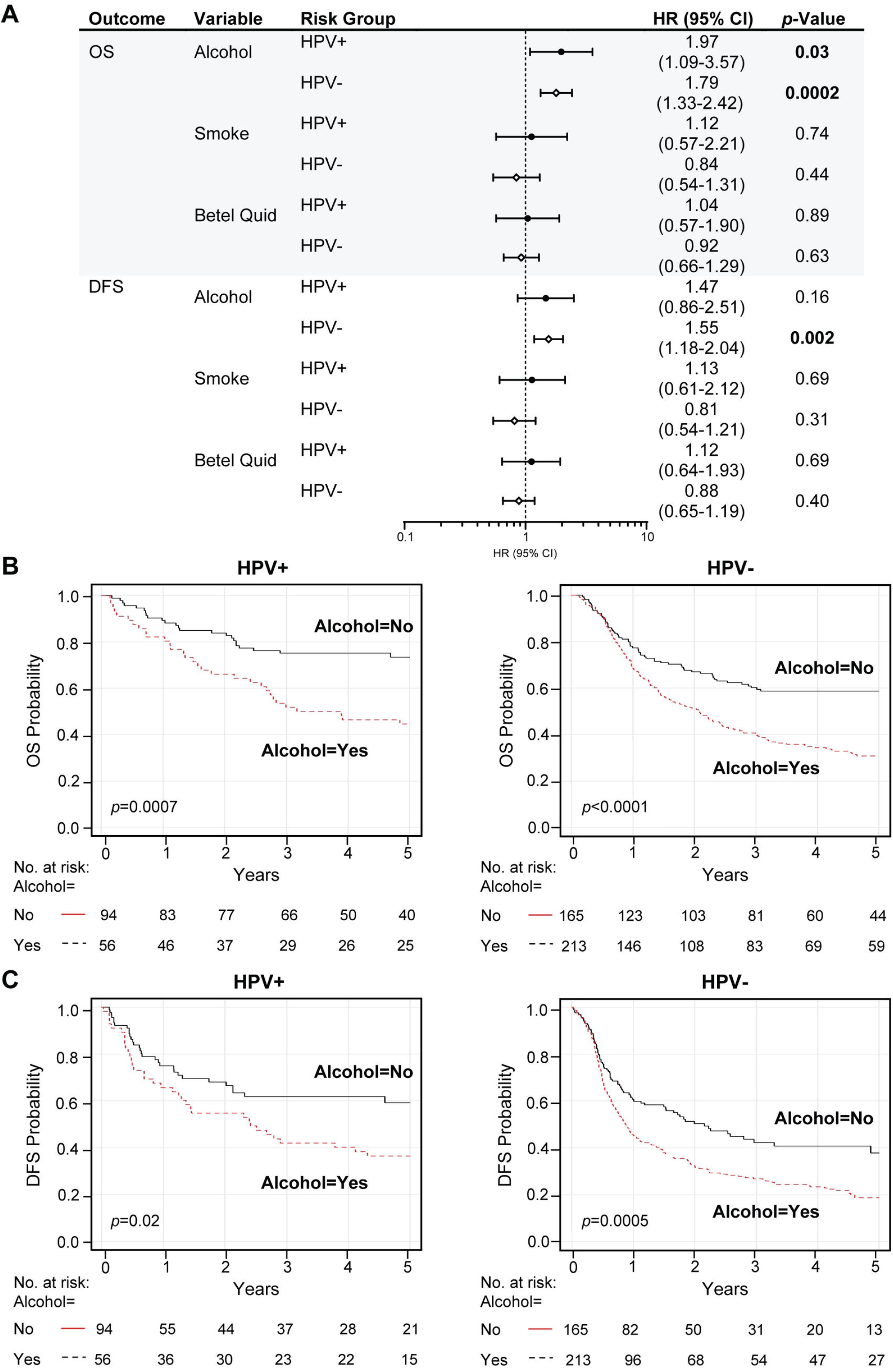
In alcohol users, HPV is associated with improved OPSCC survival time. (A-C) Prognostic outcomes of the alcohol, smoking, and betel quid variables within HPV risk groups. HPV positivity is defined as HPV DNA-positive and/or p16-positive. We analyzed up to 5-year overall survival (OS) and disease-free survival (DFS) outcomes. (A) Table includes the multivariable hazard probabilities analyzed using Cox survival models and hazard ratio (HR) estimations, adjusted for age, T- and N-stage, and visualized by forest plots. (B-C) Kaplan-Meier survival analysis. Plots represent the results for up to 5-year OS (B) and DFS (C) comparison between HPV groups stratified by alcohol groups. Log-rank analysis was used to compare the survival distributions (log-rank p-values are in the plots). The complete analysis is found in S7 Table and S3 Fig. HPV-, HPV-negative; HPV+, HPV-positive.

## Discussion

In Western countries, the prevalence of HPV-driven OPSCC has been rising drastically in the last 4 decades to become one of most common head and neck cancers [2, 4, 10, 22-24, 35-37]. Those affected by OPSCC suffer great losses due to aggressive treatment, morbidity, and death [20, 28, 35, 36, 42, 43, 45, 47-50, 69-70]. Still, the extent of this disease and its public health impact are not well understood outside North America and Western Europe. In this retrospective cohort study, we performed a comprehensive investigation of the impact of HPV-driven OPSCC in a cohort from the largest cancer treatment center in Taiwan from 1998 to 2016. We found that HPV was present in 28.4% of the tumors, with a trend for incremental occurrence over time. HPV16 was the most prevalent genotype (82.8%), followed by HPV58 (7.5%), and other diverse genotypes. HPV-positive OPSCCs occurred in higher proportion in females and presented with different clinical features than their HPV-negative counterparts, including reduced engagement in risk behaviors such as alcohol drinking, cigarette smoking, and betel quid chewing. Additional outcome analysis of the entire cohort showed that HPV-positivity was associated with a notably higher survival rate. Surprisingly, only alcohol, but not smoking or betel quid, were strongly associated with a worse prognosis. The strong prognostic benefit of HPV remained present but reduced in the presence of the associated risk factors alcohol, smoking, and betel quid.

Many studies have demonstrated an increment of HPV-driven OPSCC in numerous countries, showing considerable geographical variability in the proportion of cases over time [36, 71, 72]. Recent estimates calculate the worldwide prevalence of HPV-positive OPSCC between 18% and 35.6% [24, 73]. Reports from the United States, demonstrate a marked incremental variability over the years, from 20% by 1990 to over 70% of OPSCCs being currently caused by hrHPV [2, 7, 27, 36, 38]. Studies at the University of Michigan alone have shown a prevalence of 82.3% [27]. Reports from other developed countries and Western Europe, such as the United Kingdom and Finland, have observed a similar increment in prevalence of HPV-positive OPSCC during the last decades [7, 13, 36, 37, 39-41-72]. Differently, in South-East Asia, recent studies have shown a slower trend for these increments, with the proportion of HPV-positive OPSCC varying from 0% to 34% in various populations [9, 20, 57-62]. In neighboring Hong Kong, hrHPV has been found in 20.8% of tumors [59]. Likewise, a limited number of studies from Taiwan, suggested that hrHPV is a rising etiological factor in head and neck cancer, including OPSCC [55-57]. Particularly, Chien et al. demonstrated that in the early 2000s, 12.6% of the squamous cell tonsillar carcinomas were positive for hrHPV in Taiwan [57]. Here, our study indicates that the proportion of HPV-positive OPSCCs in the cosmopolitan Taiwanese population presented an incremental trend from 1998 to 2016. With an overall prevalence of 28.4%, our results suggest similar proportions in Taiwan to those observed in 1990 in the US [38]. Given this global historical data, we anticipate that Taiwan will also have a significant marked increment going forward in the rates of HPV-related OPSCC. Although the observed prevalence is three-fold lower in our Taiwan cohort compared with Western countries, this is higher than that reported for the US Asian population (12.8%) [46]. Our results also showed high concordance between HPV-DNA and p16 surrogate marker positivity (94.9%, 423 out of 446, *F* < 0.0001, S2 Table). p16 IHC is a robust surrogate marker and predictor for HPV-caused OPSCC, with very low percentage of false negatives (4%) [68]. These, findings indicate that HPV was present in the tested specimens and a likely etiological driver of oropharyngeal carcinogenesis, suggesting that HPV is not likely a passenger virus in most of these tumors. Still, several cases had discordant results: 2.2% (12 out of 541) tumors tested p16 negative but had HPV DNA positive results, while 2.0% (11 out of 541) tumors were p16 positive but had HPV DNA negative results (S2 Table). There are potential explanations for to account for the discordant cases. The p16-negative/HPV DNA positive cases may represent tumors where HPV is an incidental inactive passenger and not a causal driver of disease, or tumors for which p16 has been inactivated by another mechanism. Possible explanations for the p16-positive/HPV DNA negative cases could be that HPV DNA is in fact present in the tumor, but is mutated in the region where amplification/detection occurs for the test, or that p16 is upregulated by a different pathway or mechanism (something other than HPV).

Of important note, our study sheds light on the causal association between HPV genotypes and OPSCC in Taiwan. It is well-recognized that certain high-risk viral genotypes are carcinogenic and highly associated with the development of OPSCC [2, 36], especially HPV16 [3, 11, 29, 42, 73, 74]. In line with the predominant worldwide prevalence of HPV16 in OPSCC, this has also been identified in the majority of cases from Southern China [59], and previous reports in tonsillar squamous cell carcinomas in Taiwan [57]. Coincidentally, in our current study, HPV16 accounted for the vast majority (82.8%) of HPV-positive OPSCCs, followed by HPV58 in 7.5% of cases. Apart from HPV16 and HPV58, other oncogenic hrHPV genotypes were infrequent, and HPV18, which often causes cancer in the Western world, was only present in a very small proportion of the specimens (1.5% alone and 1.5% in combination with HPV16). Interestingly, our previous work has shown similar low proportion of HPV18-positive OPSCCs in the United States [65]. In addition, it has been reported that oral HPV infections in smokers and betel nut chewers, are mainly caused by HPV16. Still, HPV58 has been found at a low percentage in Northern Taiwan, especially in patients with the highest exposure to traditional habits that increase cancer risk [56]. Likewise, a literature search for the genotype prevalence of other HPV-related cancers within East Asia and Taiwan, showed that in Northern China HPV16 is dominant in cervical carcinomas [75, 76]. This prevalence decreases in the Southern regions of the country [20, 77], to become similar to the observed occurrence of other important genotypes in Taiwan, HPV52 and HPV58 [78]. Although HPV16 has been found in over 80% of cervical cancers in Asia, it has been reported in only 50% of the cervical cancers in Taiwan [79]. In this population, HPV58 is the second or the third most common genotype found in cervical malignancies (∼20%, together with HPV18) and cervical HPV infections, but it is rare in other parts of the world [80-83]. HPV52, usually found in cervical cancers in East Asia [59, 84, 85] was included in the test panel, but was not detected in our Taiwanese cohort. This particular distribution of HPV genotypes suggests tissue or geographic specificity for HPV16 and HPV58 in Taiwan, where HPV is becoming an increasingly important etiologic factor of OPSCC.

As part of our study, we also analyzed the clinical determinants of HPV positivity in OPSCC. This malignancy is considered one of the emerging causes of cancer death in Asian and Taiwan males [9, 55]. Curiously, even when most of the cases in our cohort were males (94%), females had most commonly HPV-positive tumors and presented reduced alcohol, smoking, or betel quid habits. There was also a trend for slightly better prognosis within the female group (S2 Fig). While the better prognosis in women may be related to better health utilization of women over men, our results indicate that OPSCC is a disease that also affects Taiwanese women, and more research is necessary to address their specific clinical management. Moreover, HPV-positive tumors were associated with a slightly higher age at diagnosis (mean = 55.5 years) than HPV-negative tumors. This represents another particularity since hrHPV has been historically associated with the onset of OPSCCs in younger, middle-aged individuals [23, 35, 39, 44, 59, 86], even in Taiwan, where the average age at diagnosis has been reported between 40 and 50 years of age [55]. Despite this belief, new studies from Western countries have contradicted these trends by demonstrating that the highest HPV prevalence occurs in patients above 55 and even 70 years of age [19, 87, 88]. This suggests a similar changing epidemiology in Taiwan, with a possible shift of sexual onset at a later age, reduced number of sexual partners in life, or reinfection later in life. Even when our data diverge from previous reports, these agree with studies from Southern China and Western countries indicating a significant correlation between HPV status and earlier primary tumor stage (T-stage) [3, 59]. However, our results demonstrate a higher T-stage within the entire cohort (T3-4 instead of T1-2), although the most common T class among HPV-positive tumors was T2 (46%) but among HPV-negative tumors the most common T-class was T4 (37%). N-stage did not present a significant association with HPV status. These HPV-positive tumors were associated with the tonsils as their primary location, denoting site-specificity, as also indicated in previous observations from Taiwan [55]. Thus, our results indicate that HPV-driven OPSCC is a clinically unique disease with distinctive features to this cohort of Taiwanese patients.

Furthermore, the carcinogenic effect of the risk factors alcohol, smoking, and betel quid has been very well characterized. These are considered the main etiological agents of HPV-negative OPSCC [6-13-15-17]. It has been proposed that the observed increasing rates of OPSCC in East Asia and Taiwan are related to the excessive and extensive use of alcohol, smoking, and betel quid (which does not contain tobacco) [9, 51-54-63]. Previous studies in Taiwan report that exposure to these agents represents a high risk for developing primarily intraoral cancer [9, 57]. For this reason, it was not surprising to observe that above two thirds of the OPSCC cases in our study tested negative for HPV and had a prominent smoking (83%), betel quid chewing (65%), or alcohol drinking (51%) history (Table 2, S4 Table). Earlier findings also showed a similar proportion of alcohol drinkers within the Taiwanese male population [9]. Interestingly, in our cohort, 87% of the individuals with OPSCC used these chemical carcinogens in combination. Only 1% of the cohort consumed alcohol or betel quid alone, and 11% only smoked (S4 Table). Although these historically causative risk factors are still very prominent in Taiwan, we observed that only alcohol consumption is a significant determinant of worse prognosis. This was surprising, as tobacco smoking has been found to be the most important predictor of unfavorable outcome and risk factor in OPSCC, as patients with mutant p53 have a reduced capacity to repair DNA [3, 25, 27, 44, 89]. The reasons for the dwarfed effect of smoking and betel quid as negative drivers remain elusive, but they may reside in the high proportion of smokers in the cohort (83%) or individuals with a complex combination of risk factors, as shown in S4 Table. This makes it impossible to separate the individual effects of the carcinogens, alcohol, tobacco, and betel quid, which could have additive effects on DNA damage and potentiate the negative impact of alcohol. Alternatively, alcohol could also represent a surrogate for lack of social support or socioeconomic disadvantage. Additionally, our findings reveal that HPV-positive tumors were less exposed to these three risk factors, which correlates with previous reports indicating that individuals with HPV-positive OPSCCs are more likely to be never or former smokers, or drinkers [25, 57, 59, 89, 90]. Nonetheless, these are common risk factors for HPV-positive OPSCC, and in Western countries, 10-30% of OPSCC tumors occur in individuals that smoke or drink [27, 91]. In this study, we observed a transformation from cancer that is derived from only smoking, betel quid, and alcohol influence, to an increasing trend of HPV-positive cases. The strong association of HPV with better outcome has been widely reported [23, 25, 27, 28, 35, 42, 44-50]. Our results, showing that HPV-positive OPSCCs have a strikingly better prognosis, agree with these and previous work from South-East Asia and Taiwan [57, 59]. Additional evidence showed that in our Taiwan cohort, even when having poorer outcomes, drinkers, smokers, and betel quid users benefited from the simultaneous presence of HPV, displaying higher OPSCC survival rates than those HPV-negative. Similar interactions have been seen before, especially for smoking or tobacco use and HPV [27, 89]. These results may have a substantial impact on the clinical management of OPSCC patients in Taiwan and their risk stratification. HPV-positive individuals could benefit from the secession of alcohol, smoking, or betel quid habits and therapy de-escalation, as it is currently tested in diverse hospital settings to reduce toxicity and post-treatment morbidity [13, 49, 50, 59, 92, 93].

To our knowledge, this is the first comprehensive study analyzing the impact of HPV-driven OPSCC in Taiwan. Our approach of matching HPV status and prevalence data to clinical features, risk behavior exposure, and clinical outcomes represents a distinctive strength of our study, adding to our understanding of an under-represented ethnic group in Western epidemiological studies. Also, double p16-HPV DNA testing provided clinical relevance to the findings, as the concordance of the results was very high. Because p16 as a surrogate marker for HPV-driven OPSCCs can account for more than 5% false positives [10, 94, 95], we did not rely solely on this test and performed DNA testing for HPV. Our study also suffers from limitations. Cases from a single center, the Chang Gung Memorial Hospital at Linkou, in Taiwan, were included in the study. However, it is the first and largest cancer center in Taiwan, providing cancer care to roughly a quarter of the country’s cancer patients which is a reasonably valid representation of the Taiwanese population. Although, limits to access were not assessed, which could introduce bias. An additional limitation is represented by the lack of both p16 and HPV-DNA results for all the specimens, as several were missing or had insufficient amount of tissue available for analysis (see Fig 1, S1 Table). The retrospective nature of our study also presented challenges to this work. We could not evaluate the impact of therapy because changes to treatment strategies occurred over time, and we were unable to follow changes in sexual behaviors that could perhaps help explain the occurrence of HPV in Taiwan. Data on risk factors were limited, as we could not retrieve the exact amounts and type of alcohol, cigarettes, or betel quid consumed, nor information on previous infections with hrHPV, or comorbidities. For the same reason, disease specific survival times could not be calculated. The AJCC TNM staging system changed to include HPV status in 2017 [96]; however, our cases were classified at diagnosis following previous guidance, and staging changes were not reflected in the reported N-status.

Future research should incorporate efforts to further characterize HPV-driven OPSCC in Taiwan. Since our cohort included a limited number of females, future studies aiming at clarifying the implication of this disease in women, who also suffer from HPV-driven cervical and genital cancers, are needed. Importantly, studies related to the collection of specific epidemiological data will be relevant to guide new public health policies. Since oral hrHPV infections precede the development of OPSCC, research on the natural history of this disease, including the prevalence of HPV genotypes, will help strengthen the current HPV vaccination efforts. The HPV vaccine was recently introduced in Taiwan nationwide, targeting only prepubescent girls. Still, our studies reflect that boys will also benefit from vaccination, as it has the potential to halt the expansion of HPV-positive OPSCC and other cancers [40, 56, 59]. HPV vaccination uptake is presumably low, and initially only the bivalent (HPV 16/18) and tetravalent (HPV 6/11/16/18) vaccines were used. Our data indicate that in this Taiwanese population HPV58 was the second most common hrHPV genotype. The nonavalent vaccine, which protects against HPV58 and other 8 HPV genotypes (HPV 6/11/16/18/31/33/45/52/58), would be the most appropriate choice for this target population based on our study. However, even that vaccine (without assuming cross reactivity) would not cover HPV35, 31, 59, and 66, which we found respectively account for 2.2%, 1.5%, 1.5%, and 1% each. New prospective studies have the potential to shed light on the risk of OPSCC within the vaccinated population and have broad public health implications for control measures. Lastly, multicenter, nationwide studies will provide an understanding of the variables leading to HPV oncogenesis in this population, improving risk definition, outcome prediction, and patient stratification. All these are necessary to provide patients with appropriate care based on their HPV status. Additional investigations into the molecular mechanism of HPV-induced OPSCC are also required to address the risk of recurrence and progression in Taiwan. Based on risk evaluation, HPV-positive OPSCC patients may be candidates for therapy de-intensification. Tailored interventions should also be designed based on longitudinal investigations of the interaction between HPV and risk behaviors, including drinking, smoking, and chewing betel quid.

In conclusion, our retrospective study provides empirical evidence on the impact of hrHPV on OPSCCs in a large cohort from Taiwan. We found that HPV is present and likely an increasing etiological factor in these Taiwanese individuals with OPSCC. These observations may represent continuous behavioral changes in Taiwan. HPV positivity is associated with significantly better outcomes. Involvement in risk habits, alcohol, cigarette, and betel quid use was still widespread, but the substantial prognostic benefit of HPV remained present. Thus, this consistent trend reflects the need for policies and sustained public health interventions aiming to improve the management and prevention of HPV-driven OPSCC in Taiwan.

## Supporting information

S1 Checklist

## Data Availability

All relevant data needed to reproduce our findings are included in this manuscript and its Supporting Information, excepting sensitive dates that could allow the identification of the patients in this study.

## Acknowledgments

We would like to thank the patients who generously contributed to the study, and the members of the Cancer Center, Chang Gung Memorial Hospital, for their invaluable help.

## Supporting information

**S1 Checklist. Strengthening the Reporting of Observational Studies in Epidemiology (STROBE) report**.

**S1 Fig. Representative p16 immunostaining for OPSCC tissue sections**. Specimens from example p16-negative (P0283) and p16-positive (P0267) tumors are displayed. p16 expression is observed as a brown nuclear and cytoplasmic coloration. Magnification, 200x.

**S2 Fig. Kaplan-Meier plots for survival outcomes by gender**. Up to 5-year overall survival (OS) and disease-free survival (DFS) outcomes were analyzed within the whole cohort by the Kaplan-Meier method and log-rank test (*p*-values).

**S3 Fig. Kaplan-Meier plots for survival outcomes by alcohol, smoking, and betel quid**. (A-C) Comparison of prognostic outcomes of (A) alcohol, (B) smoke, and (C) betel quid between the whole cohort and HPV risk groups. HPV positivity is defined as HPV DNA-positive and/or p16-positive. Up to 5-year overall survival (OS, top) and disease-free survival (DFS, bottom) probabilities were analyzed by the Kaplan-Meier method and log-rank test (*p*-values), as displayed for each risk group. HPV-, HPV-negative; HPV+, HPV-positive.

**S4 Fig. HPV-positive OPSCC is associated with increased disease-free survival time in the presence of other risk factors**. (A-C) Up to 5-year disease-free survival (DFS) prognostic outcome of the HPV variable within alcohol, and smoking and/or betel quid risk groups. HPV positivity is defined as HPV DNA-positive and/or p16-positive. The DFS smoking and betel quid variables were not analyzed individually due to low number of events. (A) Table includes the multivariable hazard probabilities analyzed using Cox survival models and hazard ratio (HR) estimations, adjusted for age, T- and N-stage, and which were visualized by forest plots. The complete analysis is found in S8 Table. (B-C) Kaplan-Meier survival analysis. Plots represent the DFS probabilities of cases stratified by HPV status within the (B) alcohol and (C) smoke and/or betel quid groups. Left, plots showing cases with alcohol or smoke and/or betel quid consumption. Right, plots showing cases without exposition to alcohol or smoke and/or betel quid. Log-rank analysis was used to compare the survival distributions (log-rank *p*-values are in the plots). HPV-, HPV-negative; HPV+, HPV-positive.

**S1 Table. HPV status and patient data**.

**S2 Table. p16 vs. HPV DNA results**.

**S3 Table. Yearly HPV occurrence**.

**S4 Table. Risk factors exposure characteristics, and differences regarding HPV status**.

**S5 Table. Clinical and demographic characteristics by gender**.

**S6 Table. Multivariable survival analysis of the whole cohort**.

**S7 Table. Multivariable survival analysis for alcohol, smoking, and quid within HPV r isk groups, adjusted for age, T- and N-stage**.

**S8 Table. Multivariable disease-free survival analysis by HPV within alcohol, and smoking and/or betel quid risk groups (controlled for alcohol, and smoking and/or betel quid). All models control for age, T- and N-stage**.

## Author Contributions

**Conceptualization:**Thomas E. Carey, Kai-Ping Chang, Heather M. Walline, Guadalupe Lorenzatti Hiles, Emily L. Bellile.

**Data Curation:** Guadalupe Lorenzatti Hiles, Emily L. Bellile, Chun-I Wang, Wei-Chen Yen, Lisa M. Pinatti, Heather M. Walline.

**Formal Analysis**: Guadalupe Lorenzatti Hiles, Emily L. Bellile, Heather M. Walline.

**Funding Acquisition:** Guadalupe Lorenzatti Hiles, Kai-Ping Chang, Thomas E. Carey, Heather M. Walline.

**Investigation:** Guadalupe Lorenzatti Hiles, Chun-I Wang, Wei-Chen Yen, Christine M. Goudsmit, Hannah L. Briggs, Trey B. Thomas, Lila Peters, Macy A. Afsari, Anna C. Morris, Nadine Jawad, Heather M. Walline.

**Project Administration:** Thomas E. Carey, Kai-Ping Chang, Heather M. Walline, Guadalupe Lorenzatti Hiles, Christine M. Goudsmit.

**Resources:** Kai-Ping Chang, Thomas E. Carey, Heather M. Walline.

**Supervision:** Kai-Ping Chang, Thomas E. Carey, Heather M. Walline.

**Validation:** Guadalupe Lorenzatti Hiles, Emily L. Bellile, Hannah L. Briggs, Macy A. Afsari, Heather M. Walline.

**Visualization:** Guadalupe Lorenzatti Hiles, Emily L. Bellile, Macy A. Afsari.

**Writing – Original Draft Preparation:** Guadalupe Lorenzatti Hiles.

**Writing – Review & Editing:** Guadalupe Lorenzatti Hiles, Kai-Ping Chang, Emily L. Bellile, Chun-I Wang, Wei-Chen Yen, Christine M. Goudsmit, Hannah L. Briggs, Trey B. Thomas, Lila Peters, Macy A. Afsari, Lisa M. Pinatti, Anna C. Morris, Nadine Jawad, Thomas E. Carey, Heather M. Walline.

## Funding

This study was funded by the University of Michigan-Chang Gung Memorial Hospital Pilot Grant (https://www.rogelcancercenter.org and https://www.cgmh.org.tw/en) to K-PC and TEC; the National Cancer Institute at the National Institutes of Health (https://www.cancer.gov), CA194536 to TEC and HMW, and CA194536-S1 to GLH; the Chang Gung Memorial Hospital, CMRPG3H0852, CMRPG3J1251, and CORPG3G0171 to K-PC; the Taiwan Ministry of Science and Technology (https://www.most.gov.tw), MOST 108-2314-B-182A-108-MY3 to K-PC; and, funds from the University of Michigan Undergraduate Research Opportunity Program (https://lsa.umich.edu/urop) to GLH, TEC and HMW. The funders had no role in study design, data collection and analysis, decision to publish, or preparation of the manuscript.

## Competing interests

The authors have declared that no competing interests exist.

## Abbreviations

CGMH: Chang Gung Memorial Hospital
DFS: disease-free survival
EQUATOR: Enhancing the QUAlity and Transparency Of health Research Network
FFPE: formalin-fixed, paraffin-embedded
HPV: human papillomavirus
hrHPV: high-risk human papillomavirus
IHC: immunohistochemical staining
OS: overall survival
OPSCC: oropharyngeal squamous cell carcinoma
PCR-MA: multiplex PCR-MassArray
STROBE: Strengthening the Reporting of Observational Studies in Epidemiology

## Notes

### Competing Interest Statement

The authors have declared no competing interest.

### Clinical Trial

NA

### Summary of Updates

This manuscript was sent back to us because during the screening process it came as containing too much identifying information. By error, we attached an S1 Table file that still contained the ages of the individuals in our cohort. This has been solved, leaving all minimal data necessary to replicate the study (notice that age and genre are key to the results). A new S1 Table file has been attached. The cases were unidentified in Taiwan and the sample/patient IDs were not known to anyone outside the Taiwanese research team. No other researcher from the Michigan team or outside the research group had access to patient identification. Thank you for your help!

