## Supplementary figures and images for "Understanding the impact of high-risk human papillomavirus on oropharyngeal squamous cell carcinomas in Taiwan: A retrospective cohort study"

### S1 Figure

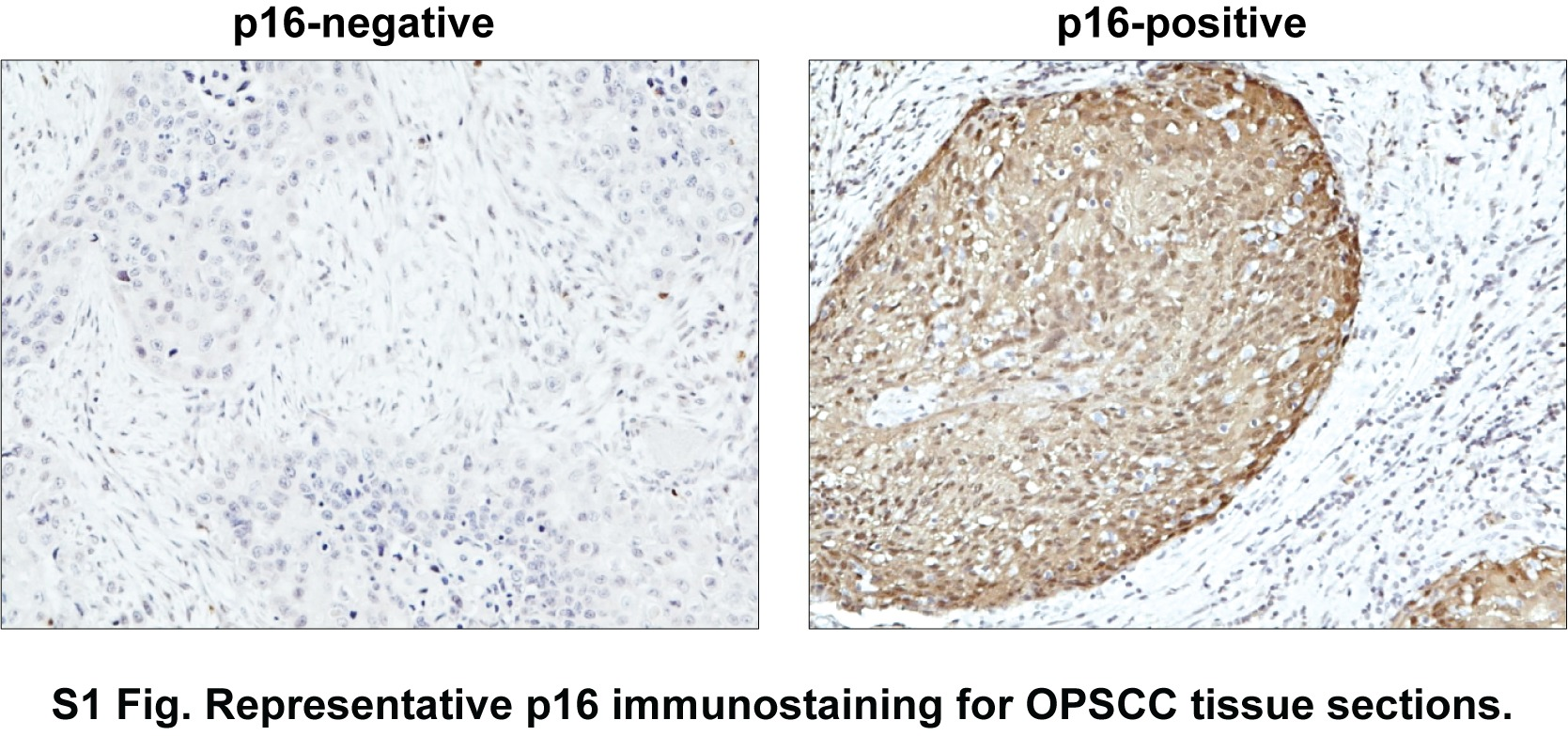

### S2 Figure

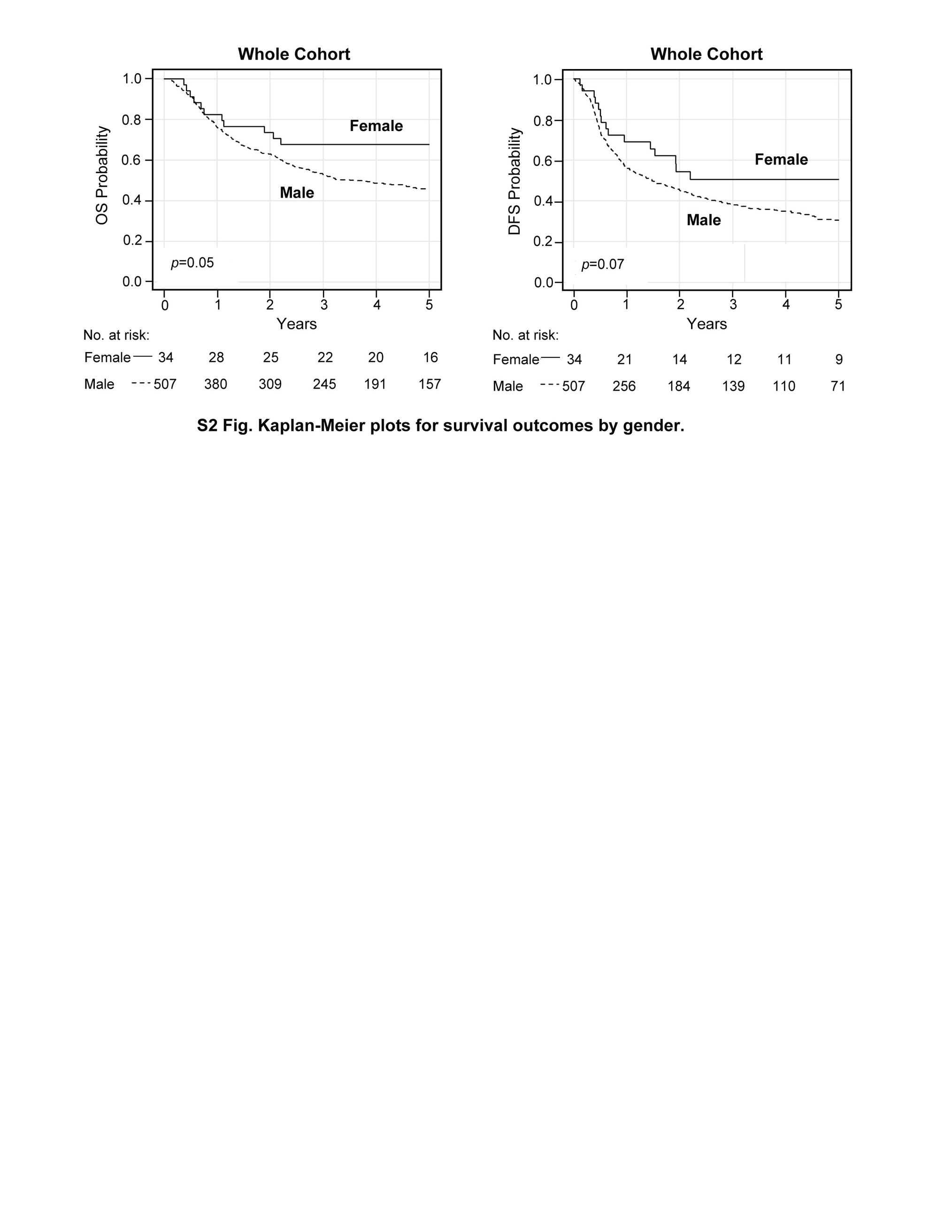

### S3 Figure

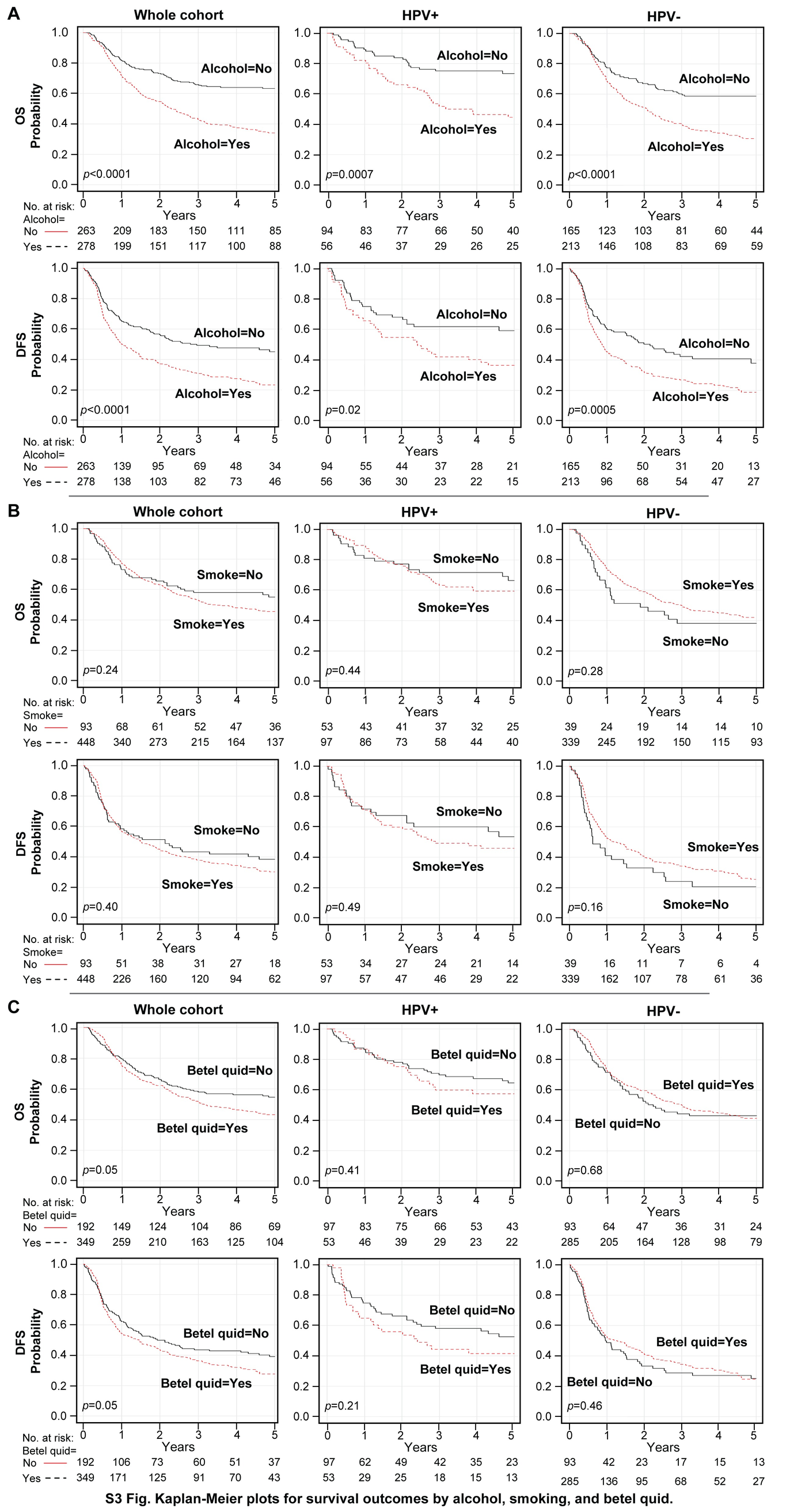

### S4 Figure

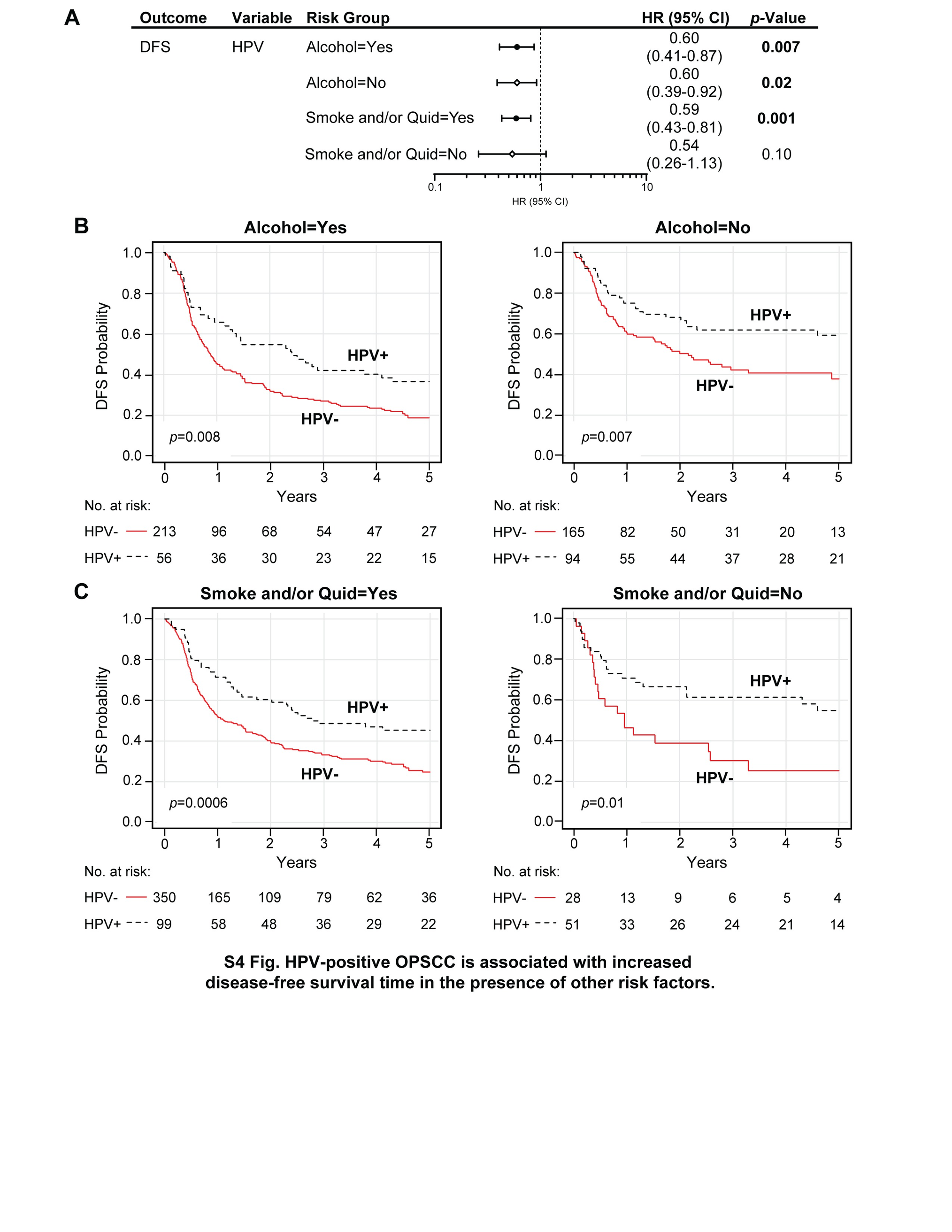
