## Supplementary material for "Understanding the impact of high-risk human papillomavirus on oropharyngeal squamous cell carcinomas in Taiwan: A retrospective cohort study": S2 Table

| **S2 Table. p16 vs. HPV DNA results.** | | | | |
| --- | --- | --- | --- | --- |
| **p16** | **HPV DNA** | | | |
|  | **. (%)** | **0 (%)** | **1 (%)** | **Total (%)** |
| **.**  **(%)** | 13  (2.40) | 35  (6.47) | 23  (4.25) | 71  (13.12) |
| **0**  **(%)** | 19  (3.51) | 324  (59.89) | 12  (2.22) | 355  (65.62) |
| **1**  **(%)** | 5  (0.92) | 11  (2.03) | 99  (18.30) | 115  (21.26) |
| **Total**  **(%)** | 37  (6.84) | 370  (68.39) | 134  (24.77) | 541  (100.00) |
| Keys. p16: (.) Slide missing or not evaluated, (0) negative, (1) positive. HPV DNA: (.) Invalid, (0) negative, (1) positive. The correlation between p16 and HPV DNA results was evaluated by Fisher's exact test (*F* < 0.0001), the corresponding values are in the central dark-gray cells. | | | | |
