## Supplementary material for "Understanding the impact of high-risk human papillomavirus on oropharyngeal squamous cell carcinomas in Taiwan: A retrospective cohort study": S3 Table

| **S3 Table. Yearly HPV occurrence.** | | | | | | | | | | | |  |  |  | |
| --- | --- | --- | --- | --- | --- | --- | --- | --- | --- | --- | --- | --- | --- | --- | --- |
| **Study Year** | **HPV-**  **(HPV DNA/p16)** | | **HPV+**  **(HPV DNA/p16)** | | **Total** | | **p16-** | | **p16+** | | **Total** | | | | |
|  | **Freq** | **%** | **Freq** | **%** | **Freq** | **%** | **Freq** | **%** | **Freq** | **%** | **Freq** | | | | **%** |
| **1998** | 14 | 58.3 | 10 | 41.7 | 24 | 100 | 15 | 62.5 | 9 | 37.5 | 24 | | | | 100 |
| **1999** | 16 | 76.2 | 5 | 23.8 | 21 | 100 | 16 | 84.2 | 3 | 15.8 | 19 | | | | 100 |
| **2000** | 21 | 67.7 | 10 | 32.3 | 31 | 100 | 19 | 79.2 | 5 | 20.8 | 24 | | | | 100 |
| **2001** | 8 | 57.1 | 6 | 42.9 | 14 | 100 | 7 | 63.6 | 4 | 36.4 | 11 | | | | 100 |
| **2002** | 17 | 65.4 | 9 | 34.6 | 26 | 100 | 16 | 69.6 | 7 | 30.4 | 23 | | | | 100 |
| **2003** | 8 | 66.7 | 4 | 33.3 | 12 | 100 | 8 | 72.7 | 3 | 27.3 | 11 | | | | 100 |
| **2004** | 16 | 94.1 | 1 | 5.9 | 17 | 100 | 15 | 100.0 | 0 | 0.0 | 15 | | | | 100 |
| **2005** | 16 | 84.2 | 3 | 15.8 | 19 | 100 | 13 | 81.3 | 3 | 18.8 | 16 | | | | 100 |
| **2006** | 15 | 100 | 0 | 0 | 15 | 100 | 12 | 100.0 | 0 | 0.0 | 12 | | | | 100 |
| **2007** | 21 | 80.8 | 5 | 19.2 | 26 | 100 | 21 | 80.8 | 5 | 19.2 | 26 | | | | 100 |
| **2008** | 30 | 68.2 | 14 | 32.8 | 44 | 100 | 24 | 75.0 | 8 | 25.0 | 32 | | | | 100 |
| **2009** | 15 | 53.6 | 13 | 46.4 | 28 | 100 | 15 | 57.7 | 11 | 42.3 | 26 | | | | 100 |
| **2010** | 35 | 83.3 | 7 | 16.7 | 42 | 100 | 30 | 81.1 | 7 | 18.9 | 37 | | | | 100 |
| **2011** | 34 | 69.4 | 15 | 30.6 | 49 | 100 | 35 | 79.5 | 9 | 20.5 | 44 | | | | 100 |
| **2012** | 30 | 65.2 | 16 | 34.8 | 46 | 100 | 25 | 61.0 | 16 | 39.0 | 41 | | | | 100 |
| **2013** | 25 | 78.1 | 7 | 21.9 | 32 | 100 | 25 | 86.2 | 4 | 13.8 | 29 | | | | 100 |
| **2014** | 31 | 79.5 | 8 | 20.5 | 39 | 100 | 30 | 78.9 | 8 | 21.1 | 38 | | | | 100 |
| **2015** | 26 | 60.5 | 17 | 39.5 | 43 | 100 | 29 | 69.0 | 13 | 31.0 | 42 | | | | 100 |
| **Total** | 378 | 71.6 | 150 | 28.4 | 528 | 100 | 355 | 75.5 | 115 | 24.5 | 470 | | | | 100 |
|  | **Frequency Missing = 13** | | | | | | **Frequency Missing = 71** | | | | | | | | |
| Study year is March-February of the following year based on the collection dates of specimens. Freq = frequency, % = percent | | | | | | | | | | | | | | | |
