## Supplementary material for "Understanding the impact of high-risk human papillomavirus on oropharyngeal squamous cell carcinomas in Taiwan: A retrospective cohort study": S4 Table

| S4 Table. Risk factors exposure characteristics, and differences regarding HPV status. | | | | | |
| --- | --- | --- | --- | --- | --- |
|  |  | **Whole cohort**  **N = 541** | **Stratified by HPV**  **N = 528** | | |
|  |  |  | **HPV-**  **N = 378** | **HPV+**  **N = 150** |  |
| Variable |  | N (%) | N (%) | N (%) | *p-*Value |
| *Risk Factors* |  |  |  |  |  |
| Smoke | **Yes** | **448 (83%)** | **339 (90%)** | **97 (65%)** | **<0.0001** |
| Betel Quid | **Yes** | **349 (65%)** | **285 (75%)** | **53 (35%)** | **<0.0001** |
| Alcohol | **Yes** | **278 (51%)** | **213 (56%)** | **56 (37%)** | **<0.0001** |
| Smoke without Alcohol and Betel Quid | **Yes** | **60 (11%)** | **33 (9%)** | **27 (18%)** | **0.0009** |
| Alcohol without Smoke and Betel Quid | **Yes** | **7 (1%)** | **1 (<1%)** | **6 (4%)** | **0.001** |
| Betel Quid without Alcohol and Smoke | **Yes** | **5 (1%)** | **3 (1%)** | **2 (1%)** | **0.5** |
| Smoke and/or Alcohol | **Yes** | **463 (86%)** | **348 (92%)** | **103 (69%)** | **<0.0001** |
| Smoke and/or Betel Quid | **Yes** | **461 (85%)** | **350 (93%)** | **99 (66%)** | **<0.0001** |
| Alcohol and/or Betel Quid | **Yes** | **408 (75%)** | **318 (74%)** | **78 (52%)** | **<0.0001** |
| Alcohol and/or Smoke and/or Betel Quid | **Yes** | **468 (87%)** | **351 (93%)** | **105 (70%)** | **<0.0001** |
| (None) | **No** | **73 (13%)** | **27 (7%)** | **45 (30%)** |  |
| HPV positivity is defined as HPV DNA-positive and/or p16-positive. *p-*values derived from Chi-square test (categorical) by HPV status, missing values were excluded. Variables were not adjusted for co-exposure to the other risk factor(s) (colored black). For individual variables “without” “Alcohol,” “Smoke,” and “Betel Quid” (colored gray), events with combined exposure to either of the other two risk factors, were removed from analysis. “Smoke and/or Alcohol” may have also used betel quid. “Smoke and/or Betel Quid” may have also used alcohol. “Alcohol and/or Betel Quid” may have also smoked. | | | | | |
