## Supplementary material for "Understanding the impact of high-risk human papillomavirus on oropharyngeal squamous cell carcinomas in Taiwan: A retrospective cohort study": S5 Table

| **S5 Table. Clinical and demographic characteristics by gender.** | | | | | |
| --- | --- | --- | --- | --- | --- |
|  |  | **Whole Cohort**  **N = 541** | **Male Only**  **N = 507** | **Female Only**  **N = 34** |  |
| **Variable** |  | N (%) | N (%) | N (%) | *p-*Value  (compared to Males) |
| **Age [Mean (std)]** | **Years** | **53.5 (10.4)** | **53.1 (10.1)** | **58.7 (12.1)** | **0.002** |
| Age | 21-40 | 42 (8%) | 42 (8%) | 0 | 0.13 |
|  | 41-60 | 358 (66%) | 336 (66%) | 22 (64%) |  |
|  | 61 to 86 | 141 (26%) | 129 (25%) | 12 (35%) |  |
| **Gender** | Male | 507 (94%) | 507 (100%) |  |  |
|  | Female | 34 (6%) |  | 34 (100%) | 0.46 |
| **Stage** | 1 | 29 (5%) | 28 (5%) | 1 (3%) | 0.32 |
|  | 2 | 68 (13%) | 64 (13%) | 4 (12%) |  |
|  | 3 | 84 (16%) | 82 (16%) | 2 (6%) |  |
|  | 4 | 359 (66%) | 332 (66%) | 27 (79%) |  |
| **T-stage** | 1 | 64 (12%) | 57 (11%) | 7 (21%) | 0.13 |
|  | 2 | 187 (35%) | 172 (34%) | 15 (44%) |  |
|  | 3 | 108 (20%) | 103 (20%) | 5 (15%) |  |
|  | 4 | 181 (34%) | 174 (34%) | 7 (21%) |  |
| **N-stage** | **0** | **169 (31%)** | **164 (32%)** | **5 (15%)** | **0.03** |
|  | **1** | **77 (14%)** | **75 (15%)** | **2 (6%)** |  |
|  | **2** | **246 (46%)** | **223 (44%)** | **23 (68%)** |  |
|  | **3** | **48 (9%)** | **44 (9%)** | **4 (12%)** |  |
| **HPV** | **HPV+** | **150 (28%)** | **129 (25%)** | **21 (62%)** | **<0.0001** |
|  | **HPV-** | **378 (72%)** | **365 (72%)** | **13 (38%)** |  |
|  | **Missing** | **13** | **13 (3%)** | **0** |  |
| **Disease Site** | **Soft Palate** | **132 (24%)** | **129 (25%)** | **3 (9%)** | **0.01** |
|  | **Tongue Base** | **90 (17%)** | **88 (17%)** | **2 (6%)** |  |
|  | **Tonsil** | **315 (58%)** | **286 (56%)** | **29 (85%)** |  |
|  | **Oropharynx, Other** | **4 (1%)** | **4 (<1%)** | **0** |  |
| **Initial Treatment** | Chemoradiation | 403 (75%) | 377 (75%) | 26 (76%) | 0.56 |
|  | Radiation | 89 (17%) | 85 (17%) | 4 (12%) |  |
|  | Surgery | 43 (8%) | 39 (8%) | 4 (12%) |  |
| ***Risk Factors*** |  |  |  |  |  |
| **Alcohol** | **Yes** | **278 (51%)** | **272 (54%)** | **6 (18%)** | **<0.0001** |
| **Smoke** | **Yes** | **448 (83%)** | **441 (87%)** | **7 (21%)** | **<0.0001** |
| **Betel Quid** | **Yes** | **349 (65%)** | **345 (68%)** | **4 (12%)** | **<0.0001** |
| ***Outcomes (within 5 years)*** | | |  |  |  |
| **Death** |  | 274 | 263 | 11 |  |
| **Recurrence** |  | 95 | 91 | 4 |  |
| **Neck recurrence** |  | 82 | 79 | 3 |  |
| **Metastasis** |  | 54 | 50 | 4 |  |
| HPV positivity is defined as HPV DNA-positive and/or p16-positive. *p-*values derived from t-test (continuous measures) or Chi-square test (categorical) by HPV status, missing values were excluded. TNM classification, according to the 7^th^ AJCC staging edition: "T" (T classification), "N" (N classification), and "Stage" (overall stage). Because all cases were presented with no metastasis (M0), there is no heading for M. | | | | | |
