## Supplementary material for "Understanding the impact of high-risk human papillomavirus on oropharyngeal squamous cell carcinomas in Taiwan: A retrospective cohort study": S6 Table

| **S6 Table. Multivariable survival analysis of the whole cohort.** | | | | | | |
| --- | --- | --- | --- | --- | --- | --- |
| **Outcome** | **Variable** | **Value** | ***p-*Value** | **HR** | **HR 95% CI** | |
|  |  |  |  |  | **Lower** | **Upper** |
| **OS** | **HPV** | **HPV+** | **0.001** | **0.58** | **0.42** | **0.81** |
|  |  | **HPV-** | **ref** |  |  |  |
|  | **Age** | 21-40 | 0.10 | 1.44 | 0.93 | 2.22 |
|  |  | **41-60** | **ref** |  |  |  |
|  |  | **61-86** | **0.01** | **1.45** | **1.08** | **1.93** |
|  | **T-stage** | **T1** | **<0.0001** | **0.33** | **0.21** | **0.54** |
|  |  | **T2** | **<0.0001** | **0.42** | **0.31** | **0.54** |
|  |  | **T3** | **0.002** | **0.59** | **0.42** | **0.82** |
|  |  | **T4** | **ref** |  |  |  |
|  | **N-stage** | **N0** | **ref** |  |  |  |
|  |  | N1 | 0.77 | 1.06 | 0.71 | 1.59 |
|  |  | N2 | 0.54 | 1.10 | 0.81 | 1.49 |
|  |  | **N3** | **0.0006** | **2.09** | **1.37** | **3.17** |
|  | **Alcohol** | **Y** | **<0.0001** | **2.06** | **1.54** | **2.74** |
|  |  | **N** | **ref** |  |  |  |
|  | Smoke | Y | 0.18 | 0.76 | 0.50 | 1.14 |
|  |  | N | ref |  |  |  |
|  | Betel Quid | Y | 0.60 | 0.92 | 0.67 | 1.27 |
|  |  | N | ref |  |  |  |
| **DFS** | **HPV** | **HPV+** | **<0.0001** | **0.54** | **0.40** | **0.73** |
|  |  | **HPV-** | **ref** |  |  |  |
|  | Age | 21-40 | 0.76 | 1.07 | 0.71 | 1.61 |
|  |  | 41-60 | ref |  |  |  |
|  |  | 61-86 | 0.20 | 1.20 | 0.91 | 1.57 |
|  | **T-stage** | **T1** | **0.0002** | **0.46** | **0.31** | **0.69** |
|  |  | **T2** | **<0.0001** | **0.54** | **0.41** | **0.71** |
|  |  | **T3** | **0.04** | **0.72** | **0.53** | **0.98** |
|  |  | **T4** | **ref** |  |  |  |
|  | **N-stage** | **N0** | **ref** |  |  |  |
|  |  | N1 | 0.95 | 0.99 | 0.69 | 1.42 |
|  |  | N2 | 0.53 | 1.09 | 0.83 | 1.44 |
|  |  | **N3** | **0.003** | **1.83** | **1.23** | **2.71** |
|  | **Alcohol** | **Y** | **<0.0001** | **1.72** | **1.33** | **2.24** |
|  |  | **N** | **ref** |  |  |  |
|  | Smoke | Y | 0.26 | 0.81 | 0.56 | 1.17 |
|  |  | N | ref |  |  |  |
|  | Betel Quid | Y | 0.46 | 0.90 | 0.66 | 1.21 |
|  |  | N | ref |  |  |  |
| Multivariable Cox proportional hazard regression analysis of each outcome. Estimates reported for full model with all covariates included as fixed effects. (R3) HPV positivity is defined as HPV DNA-positive and/or p16-positive. OS, overall survival; DFS, disease-free survival. Ref = reference category. | | | | | | |
