## Supplementary material for "Understanding the impact of high-risk human papillomavirus on oropharyngeal squamous cell carcinomas in Taiwan: A retrospective cohort study": S7 Table

| **S7 Table. Multivariable survival analysis for alcohol, smoking, and quid within HPV risk groups, adjusted for age, T- and N-stage.** | | | | | | | | | | |
| --- | --- | --- | --- | --- | --- | --- | --- | --- | --- | --- |
|  |  |  | **HPV-positive** | | | | **HPV-negative** | | | |
| **Outcome** | **Variable** | **Value** | ***p*-Value** | **HR** | **HR 95% CI** | | ***p*-Value** | **HR** | **HR 95% CI** | |
|  |  |  |  |  | **Lower** | **Upper** |  |  | **Lower** | **Upper** |
| **OS** | Alcohol | Y | **0.03** | **1.97** | **1.09** | **3.57** | **0.0002** | **1.79** | **1.33** | **2.42** |
|  |  | N |  | **ref** |  |  |  | **ref** |  |  |
|  | Smoke | Y | 0.74 | 1.12 | 0.57 | 2.21 | 0.44 | 0.84 | 0.54 | 1.31 |
|  |  | N |  | ref |  |  |  | ref |  |  |
|  | Betel Quid | Y | 0.89 | 1.04 | 0.57 | 1.90 | 0.63 | 0.92 | 0.66 | 1.29 |
|  |  | N |  | ref |  |  |  | ref |  |  |
| **DFS** | Alcohol | Y | 0.16 | 1.47 | 0.86 | 2.51 | **0.002** | **1.55** | **1.18** | **2.04** |
|  |  | N |  | ref |  |  |  | **ref** |  |  |
|  | Smoke | Y | 0.69 | 1.13 | 0.61 | 2.12 | 0.31 | 0.81 | 0.54 | 1.21 |
|  |  | N |  | ref |  |  |  | ref |  |  |
|  | Betel Quid | Y | 0.69 | 1.12 | 0.64 | 1.93 | 0.40 | 0.88 | 0.65 | 1.19 |
|  |  | N |  | ref |  |  |  | ref |  |  |
| Cox proportional hazard regression analysis of each outcome within each HPV status cohort. Behavior variable added individually to model adjusted for age, T- and N-stage. HPV positivity is defined as HPV DNA-positive and/or p16-positive. OS, overall survival; DFS, disease-free survival. Ref = reference category. | | | | | | | | | | |
