## Supplementary material for "Understanding the impact of high-risk human papillomavirus on oropharyngeal squamous cell carcinomas in Taiwan: A retrospective cohort study": S8 Table

| **S8 Table. Multivariable disease-free survival analysis of HPV status within alcohol, and smoking and/or betel quid risk groups (stratified by alcohol, and smoking and/or betel quid). All models control for age, T- and N-stage.** | | | | | | | | | | |
| --- | --- | --- | --- | --- | --- | --- | --- | --- | --- | --- |
|  |  |  | **Alcohol = Yes** | | | | **Alcohol = No** | | | |
| **Outcome** | **Variable** | **Value** | ***p*-Value** | **HR** | **HR 95% CI** | | ***p*-Value** | **HR** | **HR 95% CI** | |
|  |  |  |  |  | **Lower** | **Upper** |  |  | **Lower** | **Upper** |
| **DFS** | HPV | HPV+ | **0.007** | **0.60** | **0.41** | **0.87** | **0.02** | **0.60** | **0.39** | **0.92** |
|  |  | HPV- |  | **ref** |  |  |  | **ref** |  |  |
|  |  |  | **Smoke and/or Quid = Yes** | | | | **Smoke and/or Quid = No** | | | |
| **Outcome** | **Variable** | **Value** | ***p*-value** | **HR** | **HR 95% CI** | | ***p*-value** | **HR** | **HR 95% CI** | |
|  |  |  |  |  | **Lower** | **Upper** |  |  | **Lower** | **Upper** |
| **DFS** | HPV | HPV+ | **0.001** | **0.59** | **0.43** | **0.81** | 0.10 | 0.54 | 0.26 | 1.13 |
|  |  | HPV- |  | **ref** |  |  |  | ref |  |  |
| Cox proportional hazard regression analysis of each outcome in each risk group. HPV variable added individually to model adjusted for age, T- and N-stage. HPV positivity is defined as HPV DNA-positive and/or p16-positive. DFS, disease-free survival. Ref = reference category. | | | | | | | | | | |
